# Lysosomal enzyme deficiency and *GBA* mutations in Dystonia

**DOI:** 10.1101/2020.08.27.20182667

**Authors:** Sebastian R. Schreglmann, Derek Burke, Amit Batla, Nikola Kresojevic, Nicholas Wood, Simon Heales, Kailash P. Bhatia

**Affiliations:** Institute of Neurology, UCL, 33 Queen Square, London WC1N 3BG, UK; Enzyme Unit, Great Ormond Street Hospital, London WC1N 3JH; Department of Neurology, Clinical Centre of Serbia, 11000 Belgrade, Serbia; UCL BRC Great Ormond Street Institute of Child Health

**Keywords:** Glucocerebrosidase, pathophysiology, cerebellum, dentate nucleus

## Abstract

Glucocerebrosidase (GCase) deficiency due to mutations of the glucosidase acid beta (*GBA*) gene causes autosomal-recessive Gaucher’s disease, the most frequent lysosomal storage disorder. Over the past two decades, *GBA* mutations have been established as the most frequent genetic risk factor to develop Parkinson’s Disease. In dystonia, the underlying aetiology in a relevant proportion of cases remains unknown, hampering the development of causative treatment strategies. Here, we explored the possible role of lysosomal dysfunction in clinical (n=130) and post mortem (n=10) patients with dystonia.

As part of extensive diagnostic evaluations (screening for structural, acquired and degenerative causes of dystonia), lysosomal enzyme activity was measured in n=79 retrospectively collected cases of patients with combined dystonia and n=51 prospectively collected cases of patients with cervical dystonia using a clinically validated, fluorescence-based assay. Clinical information on all cases was extensively reviewed and an alternative aetiology of dystonia was identified in n=14 cases on follow-up. Of the remaining n=116 cases of dystonia of unknown origin, complete Sanger Sequencing of *GBA* exons 1-11 was performed using an established protocol in all n=97 of cases with available DNA. Where there was suspicion based on clinical examination or family history, nigro-striatal degeneration was excluded in n=19 (17.2%) cases with dystonia of unknown origin. Furthermore, lysosomal enzyme activity was measured in different brain regions of age-, sex- and post-mortem delay-matched cases with dystonia of unknown origin (n=10) and healthy controls (n=10) from the Queen Square brain bank.

Among cases with dystonia of unknown origin, decreased white cell Glucocerebrosidase activity was measured in a range typical for homozygous (n=2; 1.7%) or heterozygous (n=23; 19.8%) GBA mutation carriers. The frequency of *GBA* mutations (5/80=6.25%) was significantly higher in patients than in controls (3/257=1.17%) of a historical control group from the same ethnic background (P=0.02; Odds Ratio=5.64, 95% Confidence Interval=1.44 – 21.58) – known pathogenic mutations E326K, T369M and N370S were found. We also identified lower Glucocerebrosidase activity in the cerebellar dentate nucleus (P=0.048) of dystonia patients than healthy controls.

This study provides evidence for peripheral and central lysosomal dysfunction in a significant proportion and across the clinical spectrum of dystonia. As in Parkinson’s disease, this was found irrespective of *GBA* mutation status, indicating a possible role of lysosomal dysfunction as a more general disease mechanism in dystonia.

## Introduction

Lysosomal storage disorders are a heterogeneous group of conditions sharing the incomplete processing and accumulation of proteins within the lysosome, leading to progressive functional impairment (Hers, 1963). Gaucher’s disease, the most frequent lysosomal storage disorder, is characterized by decreased activity of the enzyme glucocerebrosidase (GCase), causing glucosylceramide to accumulate in cells of the monocyte-macrophage system (Grabowski, 2008), presenting with a diverse (Hruska et al., 2008) and continuous clinical spectrum (Goker-Alpan et al., 2003; Mullin et al., 2019; Sidransky, 2012).

Gaucher’s disease is caused by mutations in the 7604 base pair *GBA* gene (MIM# 606463), located on chromosome 1q21 and comprising of 11 exons and 10 introns (Horowitz et al., 1989), coding for GCase (Li et al., 2015). A non-translated 5769 base-pair pseudo-gene *(GBAP)*, sharing 96% exonic sequence homology, is located together with another six genes in close proximity (Horowitz et al., 1989), which explains frequent chromosomal rearrangements and misalignments in this region (Tayebi et al., 2003; Winfield et al., 1997). Currently there are over 300 different mutations described for *GBA* (Beutler et al., 2005; Koprivica et al., 2000; Winfield et al., 1997)(for an updated list, see: http://www.hgmd.cf.ac.uk/ac/all.php). Although known to cause Gaucher’s disease, these mutations can also be found in clinically unaffected individuals in a population-dependent manner: in studies that examined the full *GBA* sequence, this frequency has been reported in 7.1% of Ashkenazy Jews (Clark et al., 2007), respectively in 0.5% of a Spanish (Setó-Salvia et al., 2012), 0.7% of a Portuguese (Bras et al., 2009), 1.0% of a French (Lesage et al., 2010), 1.17% of a British (Neumann et al., 2009) and 2.1% of a US-American Caucasian sample (Clark et al., 2007).

Over the past two decades, an association between *GBA* mutations and Parkinson’s disease increasingly became apparent (Brockmann and Berg, 2014; Neumann et al., 2009; Sidransky et al., 2009). First detected by meticulous clinical observation of parkinsonian features in individual Gaucher’s disease cases (Neil et al., 1979; Neudorfer et al., 1996), this was substantiated, among other, by a higher Parkinson’s disease conversion rate among Gaucher’s disease patients than individuals from the general population (Goker-Alpan et al., 2004) and the detection of *GBA* mutations in pathologically confirmed Parkinson’s disease cases (Lwin et al., 2004). By now, *GBA* mutations are established to constitute the most frequent risk factor to develop Parkinson’s disease (Bultron et al., 2010; McNeill et al., 2012; Sidransky et al., 2009): pathogenic *GBA* mutations have been found in up to 7% of non-Jewish Parkinson’s disease patients (Sidransky et al., 2009). Beyond Parkinson’s disease, *GBA* mutations have been associated with and carry a higher risk for Lewy body dementia (OR 2.5, 95%CI: 1.88-3.46) than ApoE (OR 2.4) or α-synuclein mutations (OR 0.73) (Guerreiro et al., 2018).

Dystonia, characterized by involuntary excessive muscle activity, often associated with pain and work impairment, can present as isolated or combined dystonia (Albanese et al., 2013). Diagnosis frequently follows established, specific dystonia syndromes that can guide the diagnostic approach, but in the majority of cases the underlying aetiology remains unknown (Jinnah and Factor, 2015). For the majority of isolated focal dystonia, for example, after exclusion of Wilson’s disease, structural abnormalities and potential iatrogenic causes, the large majority of cases remain “idiopathic” (Evatt et al., 2011). In cervical dystonia, the most frequent form of focal dystonia, the rate of idiopathic cases has been reported as high as 84% (Strader et al., 2011). Without better understanding of the underlying disease mechanism, these cases rely on symptomatic treatment only.

In this work we report the observation of decreased lysosomal GCase enzyme activity in peripheral white blood cells in a significant proportion of patients with combined as well as isolated cervical dystonia. We furthermore explore the association between *GBA* and dystonia and report an increased frequency of known pathogenic *GBA* mutations in dystonia patients in comparison to the healthy control population of same ancestry. Examining GCase activity in post-mortem brain tissue, we also found decreased activity levels in a region-specific manner in dystonia patients.

Taken together, these results suggest the presence of lysosomal dysfunction in both peripheral as well as central tissue in a relevant minority of patients, indicating a potential role of lysosomal dysfunction in the pathophysiology of dystonia.

## Materials and methods

### Clinical cohort

Dystonia patients were recruited from the movement disorder outpatient clinic at the National Hospital of Neurology, Queen Square, London. Patients with an unknown cause of dystonia without parkinsonism and a clinical presentation deviating from recognized idiopathic dystonic syndromes (Fung et al., 2013) (e.g. late onset leg predominant dystonia, isolated foot dystonia in adults, young onset generalised dystonia negative for DYT1 mutations, rapid progressive dystonia and additional myoclonus or chorea, etc.) were included in this study and screened for structural, acquired and degenerative causes of dystonia. This included retrospective, detailed review of clinical records, neuroimaging, blood tests including routine blood count, blood film, blood chemistry for liver and renal parameters, serum copper, caeruloplasmin, genetic analysis according to clinical suspicion, organic acids, amino acids and blood white cell enzyme activity as part of their clinical diagnostic work-up. In case of clinical suspicion or a positive family history suggestive of Parkinson’s disease, the integrity of the nigrostriatal system was assessed via DaT scan imaging. Furthermore, samples from consecutive patients with sporadic cervical dystonia and normal routine diagnostic tests (Fung et al., 2013) were prospectively assessed for white blood cell (wbc) enzyme activity.

DNA samples from both prospective and retrospective cohort were collected according to ethical guidelines. The study protocol was approved by the local institutional review board/ethics committee (REC 07/Q0502/2) and all participants provided written informed consent for participation.

### Brain samples

Fresh frozen brain tissue samples from individuals with dystonia and controls without neurological symptoms were identified from the Queen Square Brain Bank at the Institute of Neurology, UCL. Written informed consent for post-mortem examination and research had been obtained in accordance with routine local procedures and was approved by the local research ethics committee. At post mortem, brains were partitioned and the right hemisphere prepared for snap freezing in −80°C liquid nitrogen vapour. Tissue microdissection was performed by an experienced dissector at the Brain Bank according to standardized procedures and samples from pallidum (PALL), cerebellar dentate nucleus (CDN), cerebellar cortex (CRB), and superior colliculus (SCoL) prepared and kept frozen on dry ice throughout processing. Samples of approximately 10mg were homogenized in a hand glass homogenizer on wet ice, homogenates diluted to a concentration of 5% weight/volume using ice cold, triple distilled water and samples fast frozen in liquid nitrogen and stored at −80°C until processing.

### Lysosomal enzyme activity measurements in blood and brain

Lysosomal enzyme activity measurements were performed in blood and brain homogenate samples using a clinical routine, fluorescence-based essay, performed at the United Kingdom Accreditation service (UKAS)-accredited laboratory of the Enzyme Unit at Great Ormond Street Children’s Hospital, London (Burke et al., 2013; Gegg et al., 2012). Based on measurements from patients with confirmed *GBA* mutation status, GCase activity ranges were categorised as typical for homozygous (<2.4μmol/l/h) or heterozygous *GBA* mutation carriers (2.5-5.4μmol/l/h), heterozygous carriers/unaffected overlap (5.4-8.9μmol/l/h) or unaffected individuals (>8.9μmol/l/h). In addition to GCase, at least beta-Galactosidase (b-GAL; reference range: 130-303μmol/l/h) and/or Chitotriosidase (CHIT; reference range 0-150μmol/l/h) activity was measured in all patient samples.

### GBA sequencing and genetic analysis

*GBA* Exons 1-11 of all available samples were fully sequenced using an established Sanger Sequencing protocol (Neumann et al., 2009; Stone et al., 2000). Chromatograms were read using CodonCodeAligner, v7.0.1 software for Mac (CodonCode Corporation, Centerville, MA, USA). Analysis of sequencing reads in forward and reverse direction was performed in clean, complete reads only and all mutations confirmed by re-amplification of the individual patient DNA. Allele names follow the common nomenclature, excluding the first 39 amino acids of the leader sequence.

### Statistical analysis

After testing for normal distribution (Shapiro-Wilk test), patient demographics and enzyme activity measurements were compared using independent samples t test and analysis of covariance (ANOVA) for parametric, respectively Mann-Whitney-U and Kruskall-Wallis test for non-parametric data – dependence of enzyme activity with age was assessed using linear regression. Frequencies of genotype, positive family history and sex were compared using a one-sided Fisher’s exact test. The degree of variation between brain enzyme activity measurements between patients and controls was compared using two sample testing of the coefficient of variation (CoV), defined as the ratio of the standard deviation to the mean. All tests were considered statistically significant in case of a *P*-value < 0.05 (two-tailed), which was adjusted in case of multiple comparison. Prism 8.0 (GraphPad Inc., San Diego, CA, USA) was used for statistical analyses.

## Data Availability

Individual level data are available on request from the corresponding author.

## Results

### Clinical cohort

Overall, white cell enzyme activity was measured in n=130 patients with dystonia (see Table 1). The genetic background of the above described cohorts was predominantly Caucasian with other ethnicities representing around one fifth of cases (see Figure 1). Of these, cases with an unknown cause of dystonia and a clinical presentation deviating from recognized dystonia syndromes had been collected retrospectively (n=79), while cases with isolated, cervical dystonia had been collected prospectively (n=51). After the identification of n=14 cases with a confirmed alternative aetiology causing dystonia (genetic, structural or alternative diagnosis) on clinical follow-up (see Table 4 for details), n=116 cases with *dystonia of unknown origin* remained. Among retrospectively collected cases (n=67), the affected body region at the onset of the condition was cervical (n=14), oro-facial / laryngeal (n=11), leg (n=14), arm/hand (n=14), generalized (n=5), or segmental (n=9). Based on clinical features, there was clinical suspicion of an underlying functional origin in n=7 cases. In comparison to prospectively collected cervical dystonia cases (n=49), retrospectively collected cases were younger (P<0.0001), had a younger age at onset (P=0.001), shorter disease duration (P=0.01) and more frequently had undergone DAT scan imaging (P=0.007; see Table 1). There was no difference in sex distribution or frequency of positive family history between cohorts.

**Table 1:**
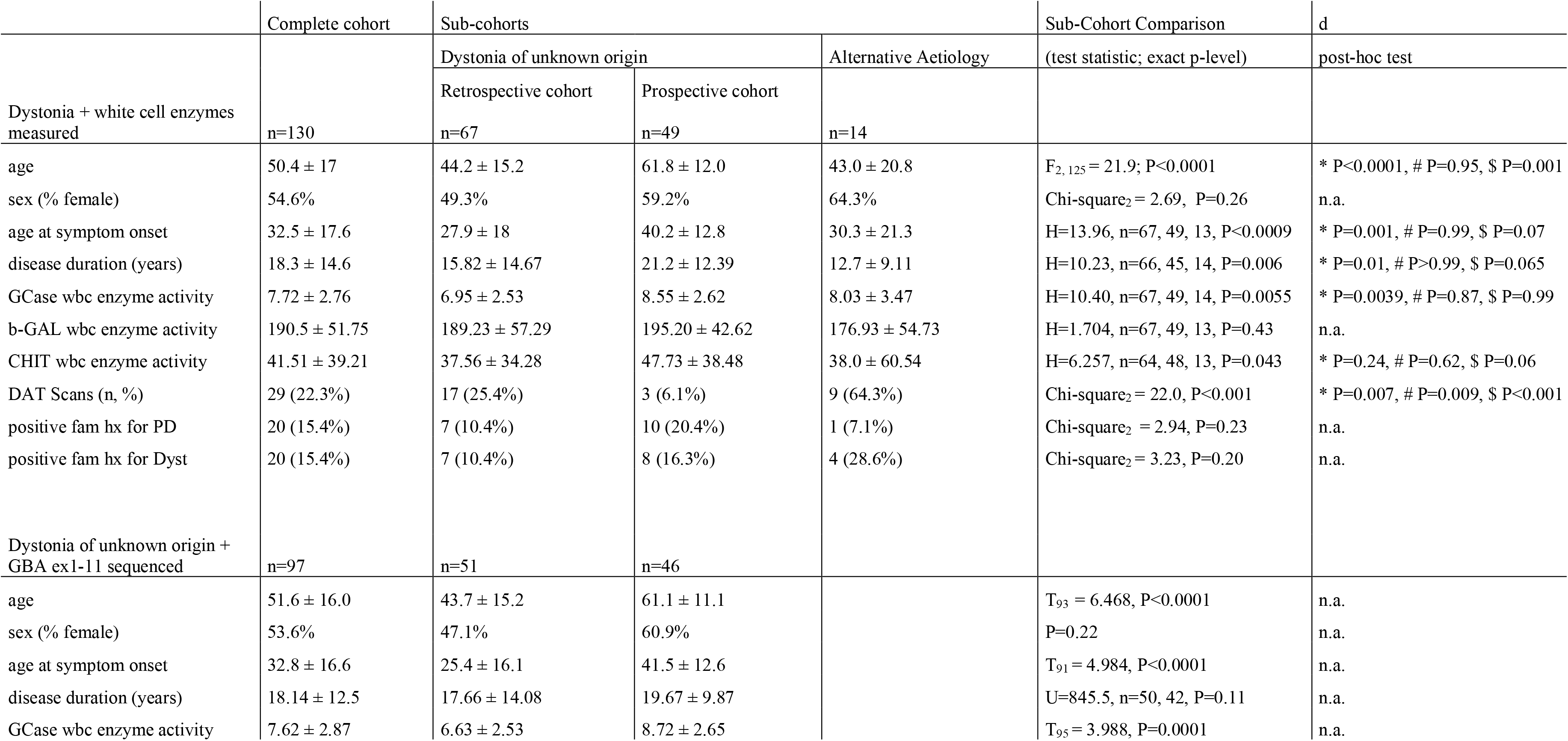
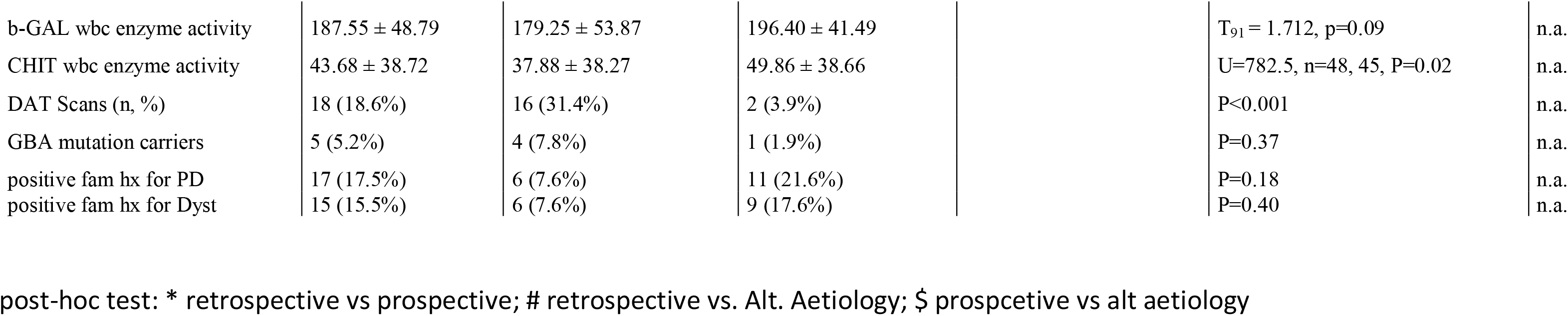
Summary clinical and demographic details of the complete cohort and each sub-cohort. (* retrospective vs prospective; # retrospective vs. alternative aetiology; $ prospective vs. alternative aetiology)

**Figure 1:**
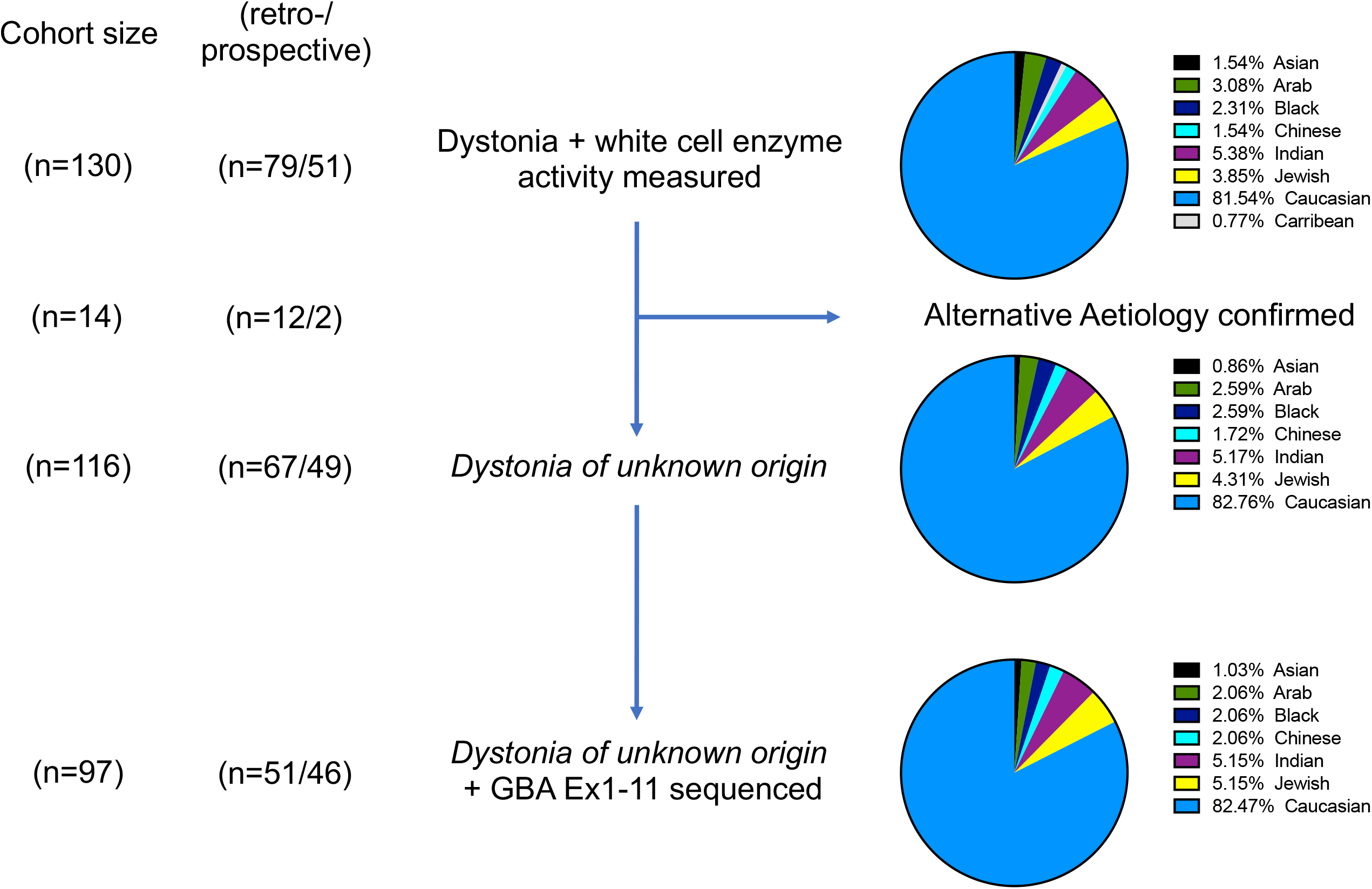
Description of the patient cohort. White blood cell lysosomal enzyme activity measurements were done in the complete cohort of n=130 dystonia patients. An alternative aetiology of dystonia was confirmed on follow-up in n=14 (see Table 4). Of the remaining n=116 patients with *dystonia of unknown origin*, DNA for complete sequencing of Exons 1-11 of the *GBA* gene was available in n=97 cases. The proportion of cases with an unknown cause of dystonia and a clinical presentation deviating from recognized dystonia syndromes (collected retrospectively) and cervical dystonia cases (collected prospectively), as well as the ethnicity distribution of respective sub-cohorts, are displayed. The proportion of patients of Caucasian ancestry did not differ between sub-cohorts.

Of the n=97 cases of *dystonia of unknown origin* for which DNA samples were available for complete Sanger sequencing of *GBA* Exons 1-11, retrospectively collected cases (n=51) were younger (P<0.0001), had a younger age at onset (P<0.0001) and more frequently had undergone DAT scan imaging (P<0.001) than prospectively collected cases (n=46). There was no difference in sex distribution, frequency of positive family history and frequency of *GBA* mutation carriers between cohorts (see Table 1).

### Lysosomal enzyme activity in blood

The activity of wbc lysosomal enzymes (in μmol/l/h) GCase, b-GAL and CHIT was compared to established, lab internal reference ranges in all n=130 cases of the combined cohort.

Among the n=116 cases of *dystonia of unknown origin*, wbc GCase activity was measured in the homozygous (n=2; 1.7%), heterozygous (n=23; 19.8%), heterozygous/overlap (n=61; 52.6%) and unaffected (n=30; 25.8%) range (Figure 2A). Mean wbc GCase activity was lower in the retrospectively vs. prospectively collected cases (P=0.0039; see Table 1). Activity of b-GAL, an indicator of general lysosomal activity, did not differ between sub-cohorts, and was found below reference range in 12 cases (retrospective n=9; prospective n=3), all of which had GCase activity within the heterozygous (n=10) or heterozygous/overlap (n=2) range. Activity in CHIT, a marker of macrophage activation, similarly did not differ between sub-cohorts, and was found mildly elevated in 3 cases (retrospective n=2; prospective n=1), all of which had GCase activity within the heterozygous/unaffected overlap range.

Among cases with *dystonia of unknown origin*, neither GCase (F_1,112_=1.308, P=0.25; r^2^=0.011) nor b-GAL (F_1,108_=0.001, P=0.97; r^2^=0.00001), but CHIT activity showed a trend to correlate with age (F_1,108_=3.726, P=0.056; r^2^=0.033; see Figure S1A-C), in keeping with the literature (Guo et al., 1995; Ramanathan et al., 2013).

**Figure 2:**
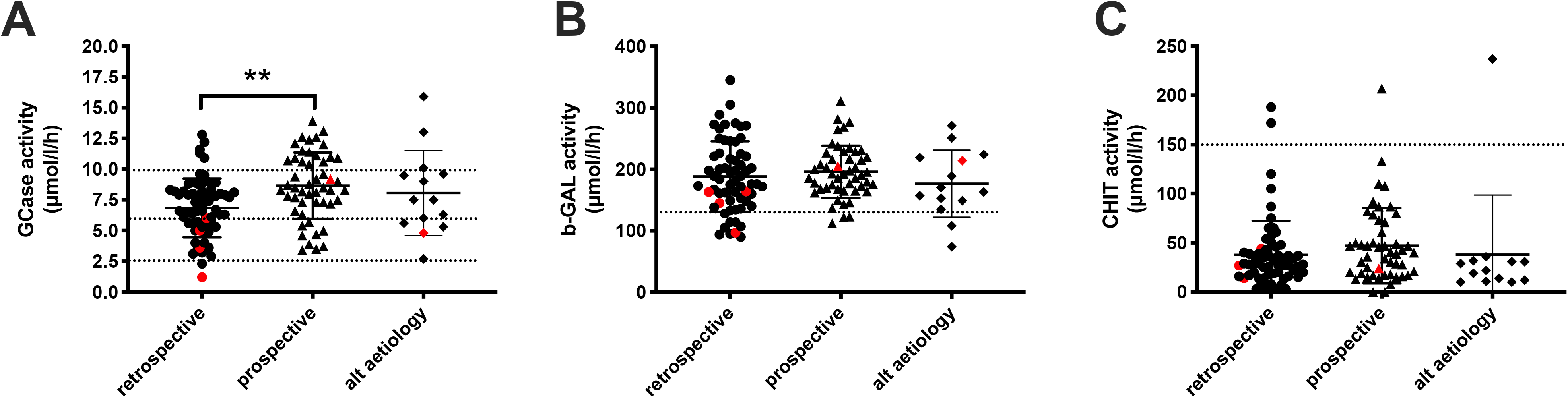
White blood cell lysosomal enzyme activity (in μmol/l/h) in dystonia patients. Enzyme activity is displayed for n=116 cases of *dystonia of unknown origin* (n=67 retrospective; n=49 prospective cases) and n=14 cases with a confirmed alternative aetiology for Glucocerebrosidase (GCase; A), beta-galactoisdase (b-GAL; B) and chitotriosidase (CHIT; C) – mean GCase activity was lower in retrospective than prospective cases (P=0.001). Dotted lines indicate internal enzyme activity reference ranges – GCase: typical for homozygous *GBA* mutations (<2.4μmol/l/h), heterozygous *GBA* mutations (2.5-5.4μmol/l/h), heterozygous / unaffected overlap (5.4-8.9μmol/l/h) or unaffected (>8.9μmol/l/h); beta-Galactosidase (b-GAL; B) normal (130-303μmol/l/h); Chitotriosidase (CHIT; C): normal (0-150μmol/l/h). Red symbols indicate cases with confirmed pathogenic *GBA* mutations.

Only mean wbc CHIT activity (male: 34.96 ± 28.24 nmol/hr/mg protein; female: 47.53 ± 41.09; Mann-Whitney U test; U=1172, P=0.026), but neither GCase activity (male: 8.01 ± 2.73; female: 7.28 ± 2.6; independent sample t test; t_114_=1,479, P=0.14) nor b-GAL activity (male: 199.0 ± 45.97; female: 185.7 ± 55.16; t_110_=1.371, P=0.17) differed between the sexes. There was a robust positive correlation between GCase and b-GAL activity (F_1,110)_=28.02, P<0.0001, r^2^=0.20, Y=8.576*X+126.7), which was not observed between GCase and CHIT (P=0.45), respectively b-GAL and CHIT (P=0.57; see Figure S1).

Similarly, in the n=97 cases of *dystonia of unknown origin* in which GBA exons 1-11 were sequenced, wbc GCase (P=0.0001) and CHIT (P=0.02), but not b-GAL activity was different between the retrospective and prospective cohort.

In the n=14 cases with a confirmed alternative aetiology, wbc GCase activity was measured in the heterozygous (n=4; 28.6%), heterozygous/overlap (n=7; 50%) and unaffected (n=3; 21.4%) range (see Table 4) and did not differ from cases with *dystonia of unknown origin* (see Table 1). GCase activity was decreased in several cases of genetically (activity level in heterozygote/overlap range in four of five cases, mean 7.2 ± 3.44μmol/l/h) and structurally confirmed alternative aetiologies (all three cases in heterozygous/unaffected overlap range, mean 6.0 ± 0.29μmol/l/h), while there was only one Parkinson’s disease case with low GCase activity, who also was found to carry a heterozygous *GBA* E326K mutation (heterozygous range; see Table 4).

### Mutations in GBA

DNA of n=97 cases of *dystonia of unknown origin* of mixed genetic background was available for sequencing. Of these, known pathogenic *GBA* mutations were found in five cases (5.15%; see Table 2). In comparison with a historic series of n=257 controls of Caucasian ancestry from the same area and sequenced according to the same sequencing protocol, which had identified three *GBA* mutation carriers (1.17%) (Neumann et al., 2009), known pathogenic *GBA* mutations were significantly more frequent among patients with *dystonia of unknown origin* (one-sided Fisher’s exact test, p=0.04; OR=4.6; 95% CI=1.18-17.57). After matching for ethnicity (five out of 80 Caucasian cases of *dystonia of unknown origin*), the frequency of *GBA* mutations in this population was 6.25% (p=0.02; OR=5.64; 95% CI=1.44-21.58).

**Table 2:** Known pathogenic *GBA* mutations identified in patients with dystonia

| Allele name<sup>a</sup> | *GBA* cDNA | Protein<sup>b</sup> | Exon | Dystonia Cases |  | Controls |  |
| --- | --- | --- | --- | --- | --- | --- | --- |
|  |  |  |  | n | Carrier frequency | n | Carrier frequency |
| E326K | c.1093G>A | p.Glu365Lys | 8 | 3 | 3% | 0 | 0% |
| T369M * | c.2023C>T | p.Thr408Met | 8 | 2 | 2% | 0 | 0% |
| N370S * | c.1226A>G | p.Asn409Ser | 9 | 1 | 1% | 1 | 0.39% |
| R257Q | c.887G>A | p.Arg296Gln | 7 | 0 | 0% | 1 | 0.39% |
| V458L | c.1489G>T | p.Val497Leu | 10 | 0 | 0% | 1 | 0.39% |
|  |  |  |  |  | 5.15% |  | 1.17% |

a Allele names follow the common nomenclature and apply to the processed protein, not including the 39-residue signal peptide.
b Protein names are based on the primary translation product and include the 39-residue signal peptide.
c Carrier frequency for each *GBA* mutation in regard to the total number of dystonia patients (total n=97).
d Carrier frequency for each *GBA* mutation in regard to the total number of historic controls from the same ethnicity and center (total n=257).
\* case 1 (see Table *GBA* mutations carriers), carried both a heterozygous T369M and heterozygous N370S variant - it was not possible to determine if in compound or combined heterozygous state (c.1226A>G(;c.2023C>T, p.Asn409Ser(;p.Thr408Met)

Three of the dystonic mutation carriers (and one case eventually diagnosed as young-onset Parkinson’s disease (YOPD)) were found to be heterozygous for E326K (carrier frequency 3%), one was heterozygous for T369M (1%), and the one with the lowest overall GCase activity carried both a heterozygous T369M and N370S mutation (1%), although it could not be determined if in combined or compound heterozygous state.

### Cases with dystonia and GBA mutations

Four of the five cases with a pathogenic *GBA* mutation belonged to the retrospectively collected group of patients with a clinical presentation deviating from recognized dystonia syndromes: e.g. Case no.1 presented with congenital cataract and isolated foot dystonia from the age of 47 and repeatedly negative DaT scans over a 14 year follow-up, Case no. 12 presented with writer’s cramp from the age of 45 developing into profound craniocervical and laryngeal dystonia, Case no.24 with dystonic posturing of both hands from the age of 15, progressive tongue biting and cervical dystonia on a background of non-REM parasomnias, Case no. 37 with the combination of upper limb dystonic posturing from the age of 13 and slowly progressive leg spasticity. There was however also a prospectively sampled case (no.99) with cervical dystonia without additional features and an inconspicuous age at onset of 44 years.

Mean age at onset among these cases was 32.4 ± 16.9 years and clinical follow-up since symptom onset was 16.6 ± 5.6 years. Wbc GCase enzyme activity was measured in the homozygous (n=1; 20%), heterozygous (n=2; 40%), heterozygous/overlap (n=1; 20%) and unaffected (n=1; 20%) range, while b-GAL was below the normal range in one case only (case no. 12).

### Cases with alternative aetiology

During clinical follow-up since initial presentation, a definitive diagnosis could be established in 14 out of 130 cases (retrospective n=12; prospective n=2; Table 4). The aetiology of dystonia was identified by either genetic testing (one each for mutations in MAPT, SCA12, SCA17, SEPN1, KMT2B n=5), additional information about previous, especially paediatric investigations (supporting pallidal necrosis, striatal lesion or ADEM during infancy; n=8) or the development of Parkinson’s disease by fulfilling clinical diagnostic criteria and showing an abnormal DaT scan (n=6).

Among these six Parkinson’s disease cases, three (cases no. 20 (carrying a heterozygous E326K *GBA* mutation), 96 & 102), developed parkinsonian symptoms within 2.3 ± 0.5 years (mean ± std.) of symptom onset and were diagnosed at 41.7 ± 6.8 years of age with YOPD. Interestingly, the other three cases (no. 105, 106 & 130) initially were diagnosed with cervical dystonia and developed parkinsonian symptoms with much more latency (14.0 ± 6.5 years) and were diagnosed at 66.3 ± 8.3 years of age as Parkinson’s disease. There were no peculiar clinical features noted in both their dystonic or parkinsonian presentation, apart from the unusually late onset of cervical dystonia symptoms in case no. 105 at 72 years of age. Only the E326K heterozygous carrier had wbc GCase activity in the heterozygous range, while in all other Parkinson’s disease cases GCase was normal.

### DaT scan imaging data

In addition to six cases that developed Parkinson’s disease on follow-up (see Table 4), DaT scan imaging data had been acquired from n=20 (17.2%) of n=116 cases with *dystonia of unknown origin*, with the majority (n=17) stemming from the retrospective cohort (see Tables 3 & 5). Case no.1 (heterozygous N370S + T369M carrier) had serial negative scans (see Supplementary Figure S2), while case no.47 was judged to have a pathological DaT scan most likely as a consequence of various lesional basal ganglia interventions decades earlier and was excluded from further analysis.

**Table 3:**
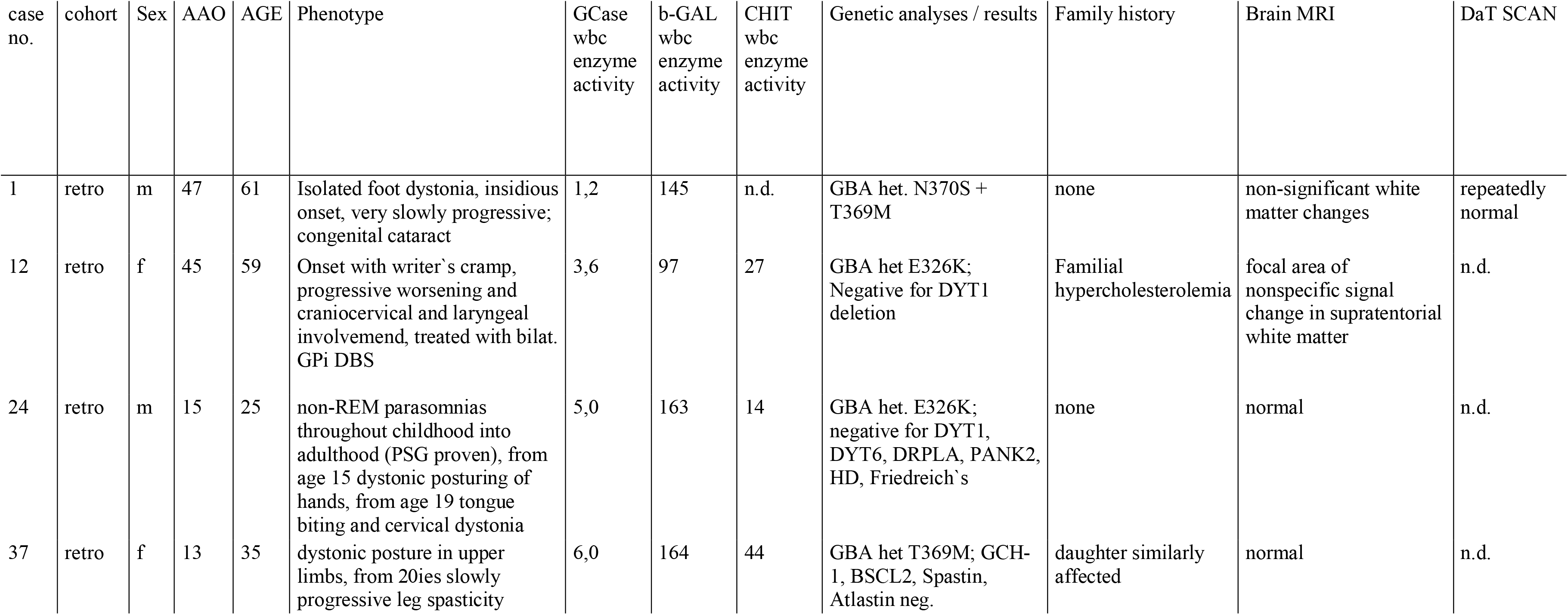
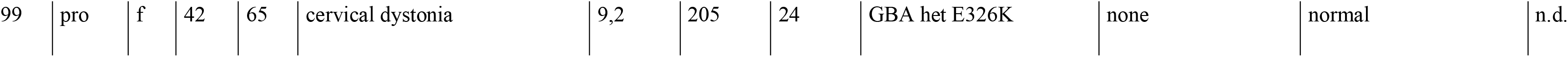
Descriptive details of dystonia patients with known pathogenic *GBA* mutations identified. All patients are of Caucasian ancestry. Lysosomal enzyme activity reference ranges: b-Glucocerebrosidase activity (homozygous: <2.4; heterozygous: 2.5-5.4; heterozygous/unaffected overlap: 5.4 – 8.9; unaffected range:8.9-16.8 μmol/l/h), b-Galactosidase activity (131-303 μmol/l/h), Chitotriosidase activity (0-150 μmol/l/h)

| case no. | cohort | Sex | AAO | AGE | Phenotype | GCase wbc enzyme activity | b-GAL wbc enzyme activity | CHIT wbc enzyme activity | Genetic analyses / results | Family history | Brain MRI | DaT SCAN |
| --- | --- | --- | --- | --- | --- | --- | --- | --- | --- | --- | --- | --- |
| 1 | retro | m | 47 | 61 | Isolated foot dystonia, insidious onset, very slowly progressive; congenital cataract | 1,2 | 145 | n.d. | GBA het. N370S + T369M | none | non-significant white matter changes | repeatedly normal |
| 12 | retro | f | 45 | 59 | Onset with writer's cramp, progressive worsening and craniocervical and laryngeal involvement, treated with bilat. GPi DBS | 3,6 | 97 | 27 | GBA het E326K; Negative for DYT1 deletion | Familial hypercholesterolemia | focal area of nonspecific signal change in supratentorial white matter | n.d. |
| 24 | retro | m | 15 | 25 | non-REM parasomnias throughout childhood into adulthood (PSG proven), from age 15 dystonic posturing of hands, from age 19 tongue biting and cervical dystonia | 5,0 | 163 | 14 | GBA het. E326K; negative for DYT1, DYT6, DRPLA, PANK2, HD, Friedreich's | none | normal | n.d. |
| 37 | retro | f | 13 | 35 | dystonic posture in upper limbs, from 20ies slowly progressive leg spasticity | 6,0 | 164 | 44 | GBA het T369M; GCH-1, BSCL2, Spastin, Atlastin neg. | daughter similarly affected | normal | n.d. |
| 99 | pro | f | 42 | 65 | cervical dystonia | 9,2 | 205 | 24 | GBA het E326K | none | normal | n.d. |

**Table 4:**
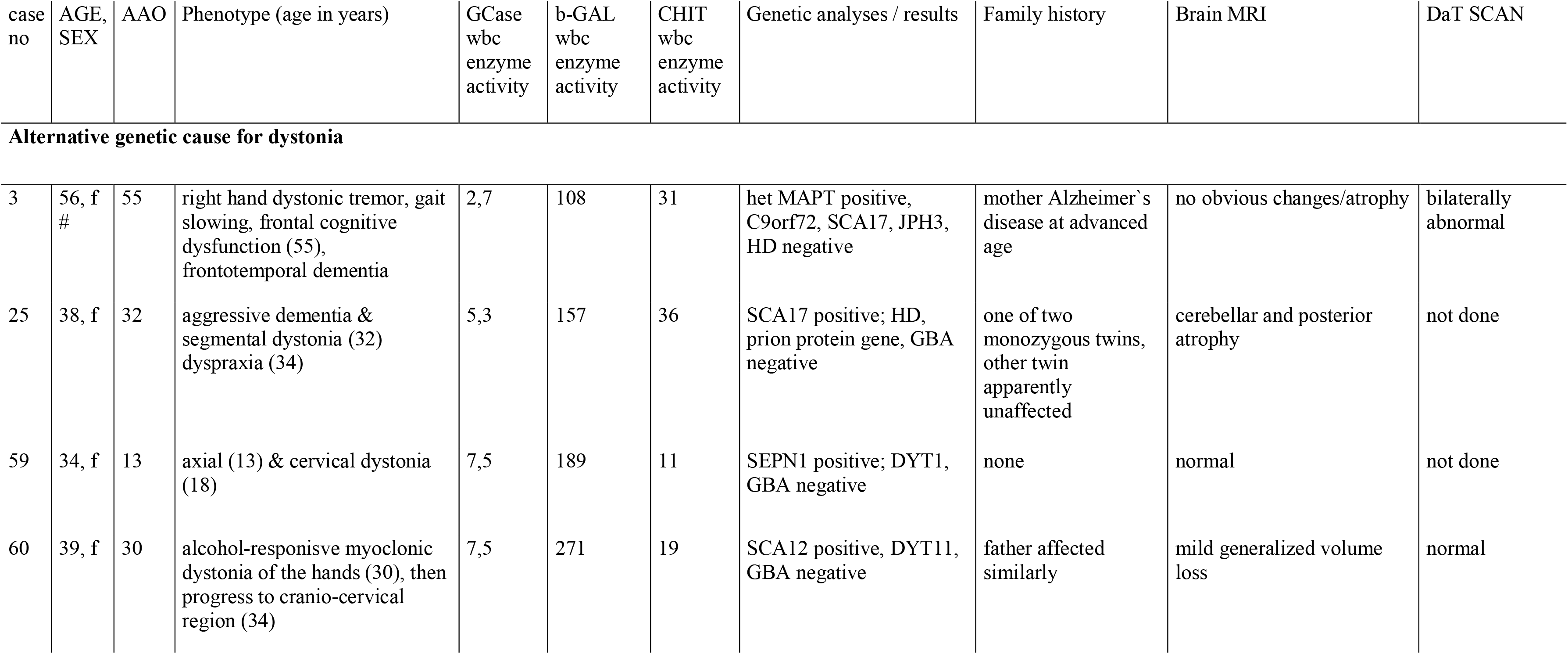
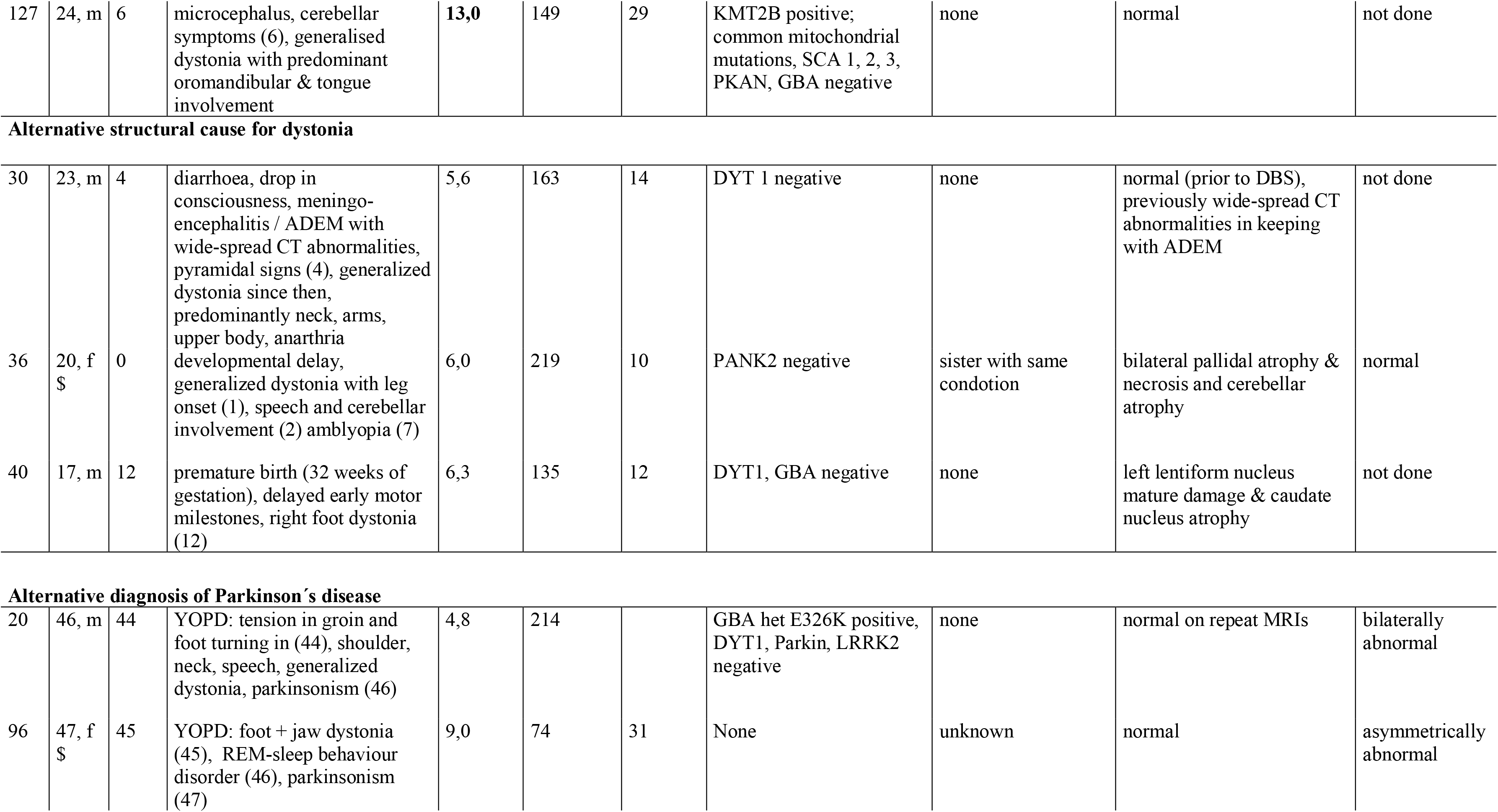
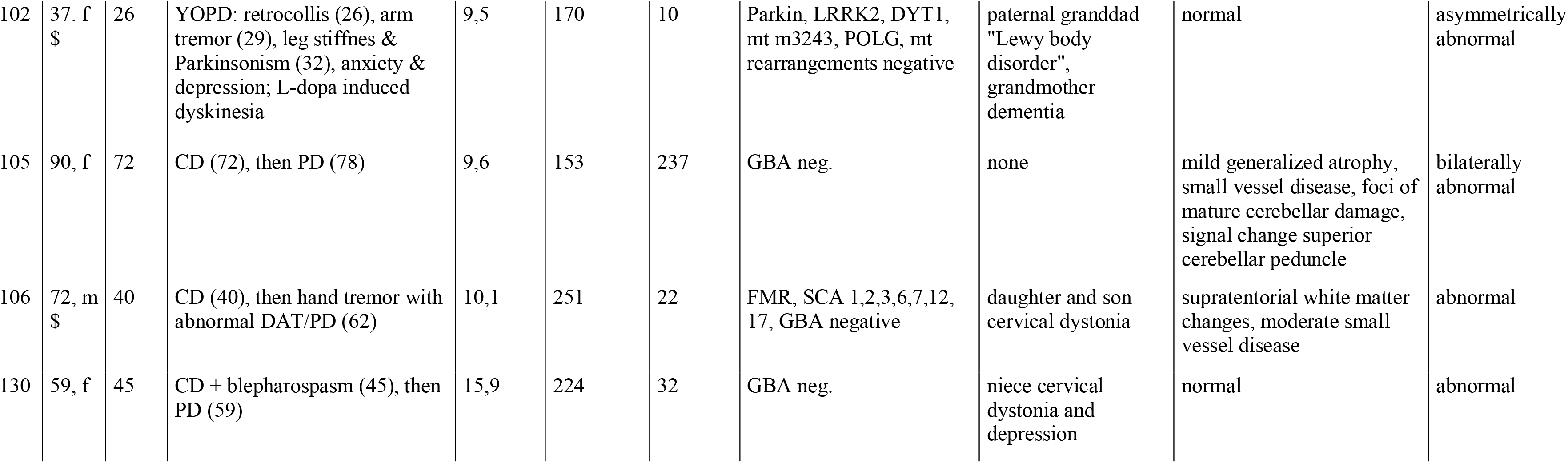
Descriptive details of patients with dystonia and confirmed alternative aetiology on clinical follow-up. heterozygous (het.), negative (neg.), # no consent for research sequencing, $ incomplete sequencing due to insufficient amount/quality of DNA sample available;

| case no | AGE, SEX | AAO | Phenotype (age in years) | GCase wbc enzyme activity | b-GAL wbc enzyme activity | CHIT wbc enzyme activity | Genetic analyses / results | Family history | Brain MRI | DaT SCAN |
| --- | --- | --- | --- | --- | --- | --- | --- | --- | --- | --- |
| **Alternative genetic cause for dystonia** |  |  |  |  |  |  |  |  |  |  |
| 3 | 56, f # | 55 | right hand dystonic tremor, gait slowing, frontal cognitive dysfunction (55), frontotemporal dementia | 2,7 | 108 | 31 | het MAPT positive, C9orf72, SCA17, JPH3, HD negative | mother Alzheimer's disease at advanced age | no obvious changes/atrophy | bilaterally abnormal |
| 25 | 38, f | 32 | aggressive dementia & segmental dystonia (32) dyspraxia (34) | 5,3 | 157 | 36 | SCA17 positive; HD, prion protein gene, GBA negative | one of two monozygous twins, other twin apparently unaffected | cerebellar and posterior atrophy | not done |
| 59 | 34, f | 13 | axial (13) & cervical dystonia (18) | 7,5 | 189 | 11 | SEPN1 positive; DYT1, GBA negative | none | normal | not done |
| 60 | 39, f | 30 | alcohol-responisve myoclonic dystonia of the hands (30), then progress to cranio-cervical region (34) | 7,5 | 271 | 19 | SCA12 positive, DYT11, GBA negative | father affected similarly | mild generalized volume loss | normal |
| 127 | 24, m | 6 | microcephalus, cerebellar symptoms (6), generalised dystonia with predominant oromandibular & tongue involvement | 13,0 | 149 | 29 | KMT2B positive; common mitochondrial mutations, SCA 1, 2, 3, PKAN, GBA negative | none | normal | not done |
| **Alternative structural cause for dystonia** |  |  |  |  |  |  |  |  |  |  |
| 30 | 23, m | 4 | diarrhoea, drop in consciousness, meningo-encephalitis / ADEM with wide-spread CT abnormalities, pyramidal signs (4), generalized dystonia since then, predominantly neck, arms, upper body, anarthria | 5,6 | 163 | 14 | DYT 1 negative | none | normal (prior to DBS), previously wide-spread CT abnormalities in keeping with ADEM | not done |
| 36 | 20, f<br>\$ | 0 | developmental delay, generalized dystonia with leg onset (1), speech and cerebellar involvement (2) amblyopia (7) | 6,0 | 219 | 10 | PANK2 negative | sister with same condotion | bilateral pallidal atrophy & necrosis and cerebellar atrophy | normal |
| 40 | 17, m | 12 | premature birth (32 weeks of gestation), delayed early motor milestones, right foot dystonia (12) | 6,3 | 135 | 12 | DYT1, GBA negative | none | left lentiform nucleus mature damage & caudate nucleus atrophy | not done |
| **Alternative diagnosis of Parkinson's disease** |  |  |  |  |  |  |  |  |  |  |
| 20 | 46, m | 44 | YOPD: tension in groin and foot turning in (44), shoulder, neck, speech, generalized dystonia, parkinsonism (46) | 4,8 | 214 |  | GBA het E326K positive, DYT1, Parkin, LRRK2 negative | none | normal on repeat MRIs | bilaterally abnormal |
| 96 | 47, f<br>\$ | 45 | YOPD: foot + jaw dystonia (45), REM-sleep behaviour disorder (46), parkinsonism (47) | 9,0 | 74 | 31 | None | unknown | normal | asymmetrically abnormal |
| 102 | 37. f<br>\$ | 26 | YOPD: retrocollis (26), arm tremor (29), leg stiffness & Parkinsonism (32), anxiety & depression; L-dopa induced dyskinesia | 9,5 | 170 | 10 | Parkin, LRRK2, DYT1, mt m3243, POLG, mt rearrangements negative | paternal granddad "Lewy body disorder", grandmother dementia | normal | asymmetrically abnormal |
| 105 | 90, f | 72 | CD (72), then PD (78) | 9,6 | 153 | 237 | GBA neg. | none | mild generalized atrophy, small vessel disease, foci of mature cerebellar damage, signal change superior cerebellar peduncle | bilaterally abnormal |
| 106 | 72, m<br>\$ | 40 | CD (40), then hand tremor with abnormal DAT/PD (62) | 10,1 | 251 | 22 | FMR, SCA 1,2,3,6,7,12, 17, GBA negative | daughter and son cervical dystonia | supratentorial white matter changes, moderate small vessel disease | abnormal |
| 130 | 59, f | 45 | CD + blepharospasm (45), then PD (59) | 15,9 | 224 | 32 | GBA neg. | niece cervical dystonia and depression | normal | abnormal |

**Table 5:**
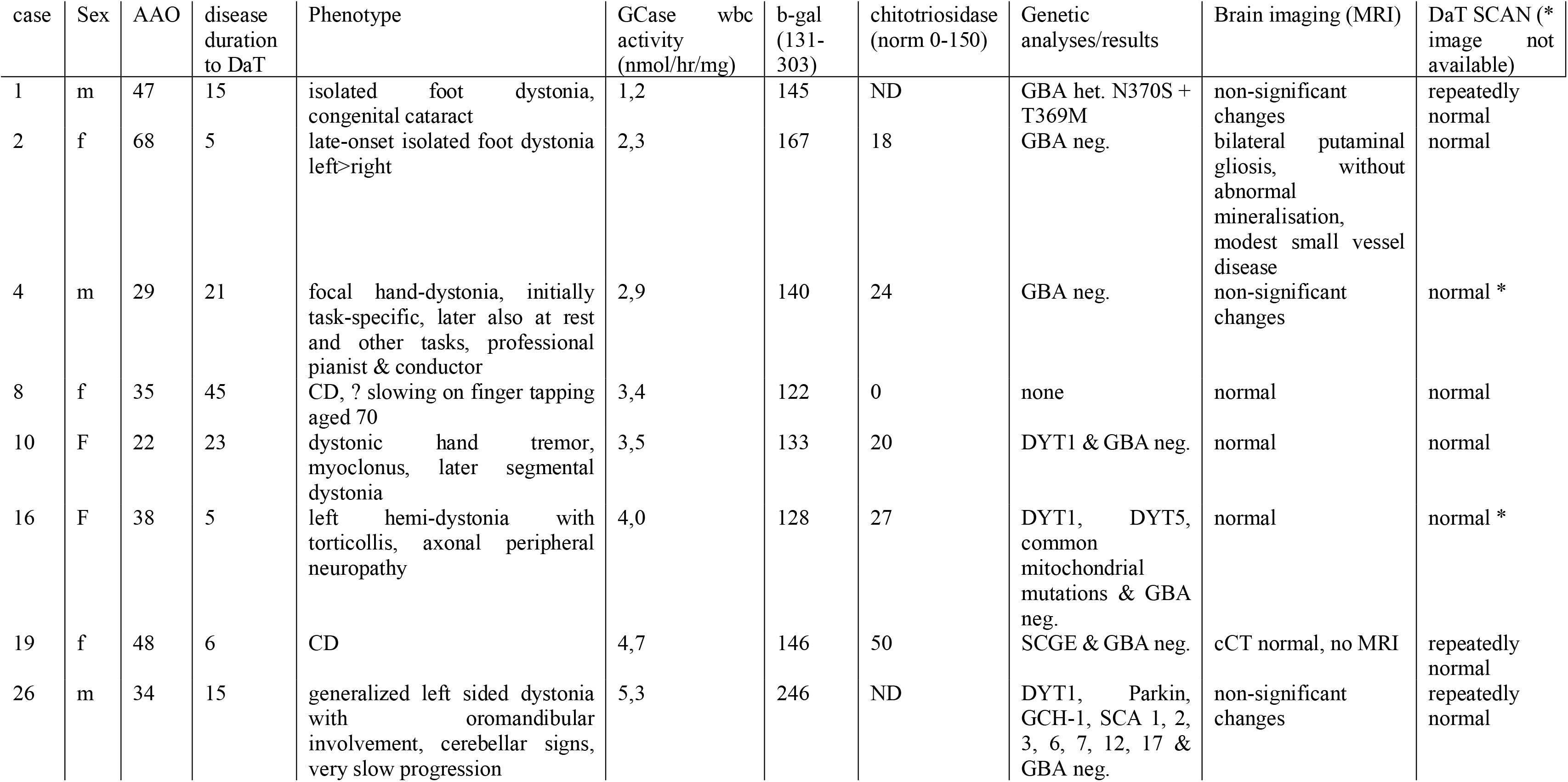
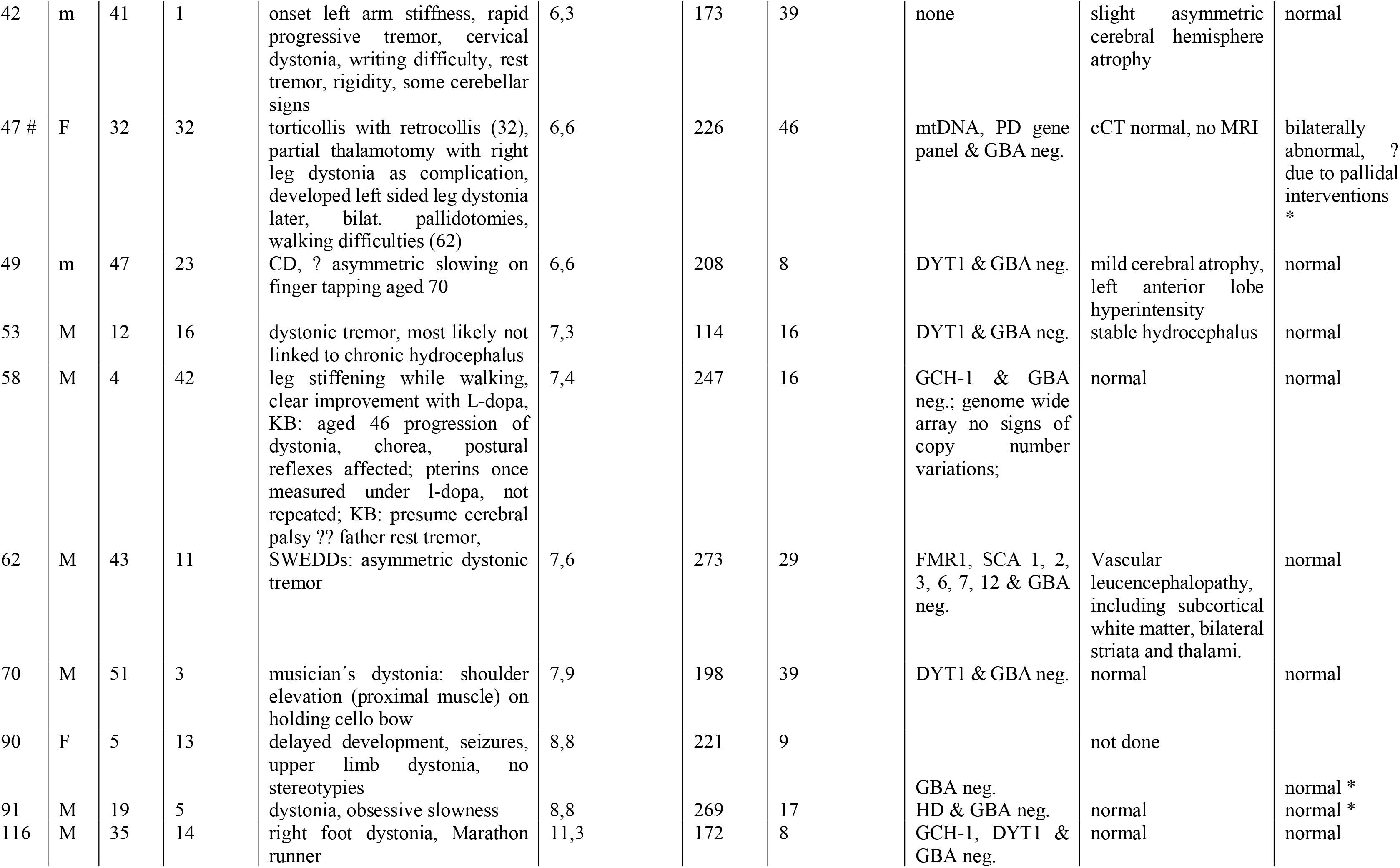
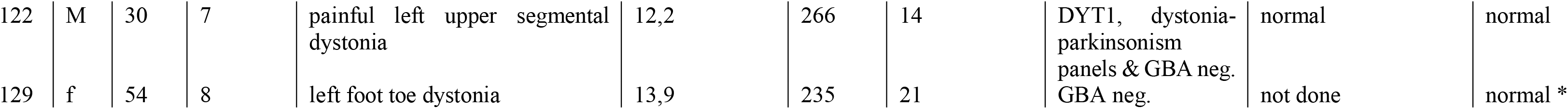
Descriptive details of patients with *dystonia of unknown origin* that underwent evaluation of the nigro-striatal system. Case no.47 (marked with #) was excluded from statistical evaluation, as imaging changes were interpreted as related to pallidotomies decades earlier.

In the remaining n=19 (16.4%) dystonia cases with normal DaT scan imaging, mean age at onset was 34.8 ± 16.5 years (range: 4 – 68 years), mean disease duration to DaT scan was 14.6 ± 11.8 years (range: 1-45 years) and wbc GCase activity was measured in the homozygous (n=2; 10%), heterozygous (n=6; 30%), heterozygous/overlap (n=8; 45%) and unaffected (n=3; 15%) range. In absolute measures, mean enzyme activity (in μmol/l/h) was 6.6 ± 3.4 for GCase, 189.6 ± 53.2 for b-GAL and 20.9 ± 12.5 for CHIT.

### Brain homogenate enzyme activity measurements

Enzyme activity (in nmol/hr/mg protein) for GCase and b-GAL were measured in brain tissue homogenates of healthy controls (Ctrl, n=10) and dystonia patients (Dystonia; n=10), matched for sex (Fisher’s exact test, p>0.99), age (two-tailed unpaired t test; t_18_=1.93, P=0.07) and post-mortem delay (t_17_=0.25, P=0.80). Due to the scarcity of dystonia donor brains, not only samples from patients with focal, segmental and generalized dystonia (n=5) but also with blepharospasm (n=5) were included.

Among Ctrl tissue samples, age at death had a significant influence on enzyme activity levels for GCase in CDN (F_1,9_=9.056, P=0.014; r^2^=0.501; Y=-0.20*X+45.42) and PALL (F_1,7_=9.807, P=0.016; r^2^=0.584; Y=0.411*X-11.28) but neither CRB (P=0.88) and SCoL (P=0.30), nor b-GAL activity in any of the regions studied (CDN, P=0.63; CRB, P=0.97; PALL, P=0.75; SCoL, P=0.35). Post mortem delay had a significant influence on enzyme activity levels for GCase in PALL (F_1,7_=11.4, P=0.011; r^2^=0.619; Y=-0.17*X+33.96), but neither for GCase (CDN, P=0.10; CRB, P=0.83; SCoL, P=0.35) nor b-GAL (CDN, P=0.09; CRB, P=0.83; PALL, P=0.52; SCoL, P=0.81) in any of the other regions studied. There was no difference between sexes in enzyme activity across brain regions. Between brain regions, GCase activity was highest in SCoL and lowest in PALL (one-way ANOVA; F_3,38_=4.336, P<0.01), whereas b-GAL activity was highest in CDN > CRB > CRB/PALL (F_3,38_=24.36, P<0.0001; see Figures 3A – B).

**Figure 3:**
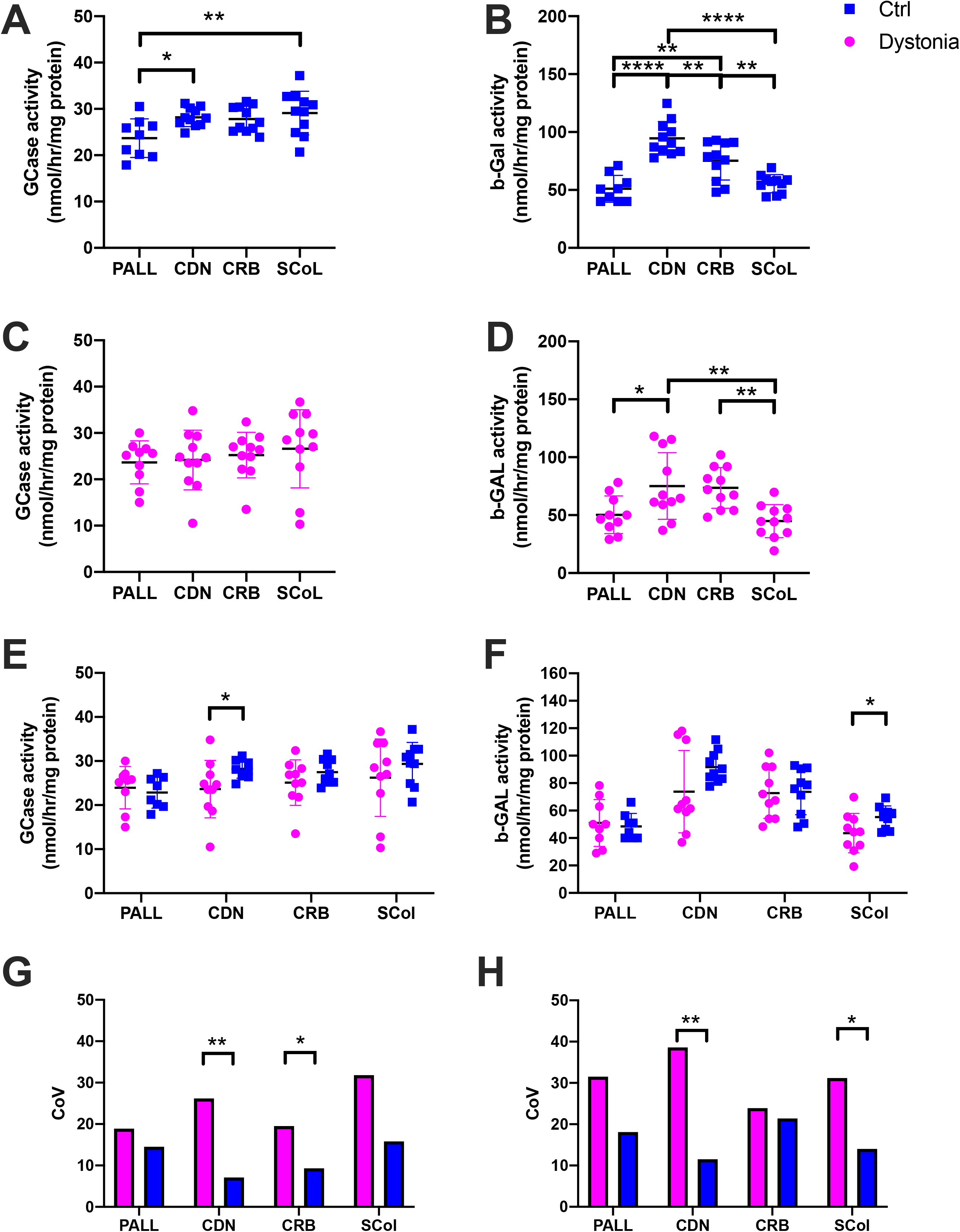
Brain homogenate lysosomal enzyme activity (in nmol/hr/mg protein) in patients with dystonia (n=10) and controls (n=10). Absolute enzyme activities (A-F) and coefficient of variation (G-H) are displayed per group and brain region. Pallidum (PALL), Cerebellar dentate nucleus (CDN), cerebellar cortex (CRB), superior colliculus (SCol);

This region-specific pattern was lost in dystonia tissue samples for GCase (F_3,39_=0.45, P=0.72) and attenuated for b-GAL (F_3,39_=6.52, P=0.001; see Figures 3C – D). Mean activity levels for GCase were lower in dystonia vs. Ctrl tissue samples in CDN (independent sample t test; t_18_=2.12, P=0.048) and b-GAL in SCoL (t_18_=2.23, P=0.038) but did not reach statistical significance in other brain regions (see Figure 3E – F). Describing the degree of dispersion of measurements per group, the CoV was significantly larger among dystonia samples for GCase activity in CDN (P=0.0008) and CRB (P=0.03) with a trend in SCol (P=0.056), as well as for b-GAL in CDN (P=0.0023) and SCoL (P=0.03; see Figures 3G – H).

## Discussion

In this work we provide first evidence of decreased lysosomal enzyme activity in both combined and isolated dystonia. Using a comprehensive approach of enzyme activity quantification in wbc and brain tissue, as well as full sequencing of the *GBA* gene in a retrospective and a prospective cohort of dystonia patients, our results imply lysosomal dysfunction in a significant minority of dystonia cases.

We base our argument first) on the largest evaluation of peripheral lysosomal enzyme activity from dystonia patients so far, showing a reduced wbc GCase activity within the range typically measured in homozygous or heterozygous *GBA* mutation carriers in a strikingly high 21.5% of cases of *dystonia of unknown origin*. The two cohorts studied represent two distinct parts, if not extreme ends, of the phenotypical spectrum within dystonia. Although the proportion of cases with decreased wbc GCase activity (homozygous or heterozygous *GBA* mutation carrier range) was higher in the retrospective cases not fitting idiopathic dystonic syndromes (26.8%) than the prospective, homogenous cervical dystonia cohort (16.3%), the finding of a in principle similar metabolic pattern in a proportion of cases in both cohorts indicates that decreased peripheral lysosomal activity might be a common abnormality in different forms of dystonia.

Apart from Gaucher’s disease, abnormal GCase activity so far has only been described in Parkinson’s disease (Alcalay et al., 2015; 2018; Atashrazm et al., 2018). In parallel to GCase activity, reduced b-GAL activity was found in the CSF and patient-derived neurons of Parkinson’s disease patients, whereas in Gaucher’s disease b-GAL activity in fibroblasts and leukocytes has been found increased (Schöndorf et al., 2014), implying that the pattern of lysosomal activity differs between Gaucher’s disease and Parkinson’s disease. The positive correlation between peripheral GCase and b-GAL activity in our sample hence shows similarity to the metabolic pattern in Parkinson’s disease and points at a more general lysosomal dysfunction in dystonia. Similarly, normal CHIT activity in the large majority of samples argues against a macrophage activation pattern in dystonia, as present in Gaucher’s disease. The higher mean CHIT activity among female vs. male patients with *dystonia of unknown origin* (P=0.026) has not been reported in healthy controls (Guo et al., 1995; Ramanathan et al., 2013), Gaucher’s or Parkinson’s disease patients (Alcalay et al., 2015).

As wbc enzyme activity was only measured consistently for GCase, b-GAL and CHIT in this study, the full pattern of lysosomal enzyme activity in dystonia remains to be established. Nevertheless, our data suggest that the pattern of decreased activity in dystonia is similar to Parkinson’s and distinct from Gaucher’s disease.

The finding of decreased lysosomal enzyme activities in cases with an alternative genetic cause for dystonia remains inconclusive, as lysosomal activity has not been reported in carriers of *MAPT* mutations and SCA17 so far. The observation of reduced LAMP-1 staining in fronto-temporal dementia due to *MAPT* however points to relative lysosomal deficiency in this condition (Bain et al., 2019). Alternatively to being caused by genotype, GCase activity might be influenced by the dystonia disease process itself, i.e. an endophenotype like in Parkinson’s disease (Gegg et al., 2012). Further studies will need to clarify this.

Second) we identified a significantly higher rate of pathogenic *GBA* mutations among dystonia cases (6.25%) than healthy controls of the same ancestry (1.17%; p=0.02). In this cohort, heterozygous mutations E326K and T369M, as well as T369M/N370S (compound or combined heterozygous state) were detected, with E326K being the most frequent. GBA-related Parkinson’s disease has been mostly described in the presence of heterozygous (Sidransky and Lopez, 2012), and only rarely in homozygous or compound heterozygous mutation state (Clark et al., 2007; Lesage et al., 2010). Alleles E326K and T369M are established risk factors for Parkinson’s disease (OR 1.71, 95% CI 1.55-1.89, respectively OR 1.74, 95%CI 1.19-2.55) (Zhang et al., 2018) but not Gaucher’s disease (Horowitz et al., 2011; Mallett et al., 2016; Pankratz et al., 2012), while N370S is a risk factor for Parkinson’s disease (OR 3.08, 95% CI 2.32-4.09) (Pankratz et al., 2012) and the most frequent cause of Gaucher’s disease across most populations (Alfonso et al., 2007; Lacerda et al., 1994).

Similar to Gaucher’s disease, certain *GBA* variants have been reported to occur more frequently with Parkinson’s disease than others. Of 13 studies that based their results on the analysis of *GBA* exons 1-11 (including n=4966 patients from China, Portugal, Greece, Japan, Britain, North-Africa, French-Canada, Europe and Korea), eight reported L444G and six N370S as the most frequent variant (Sidransky and Lopez, 2012). Regarding individuals of Caucasian background, N370S, respectively L444P accounted for 36%/21% of carriers in a Portuguese study (Bras et al., 2009), 47% /22% in a French (Lesage et al., 2010) and 24%/33% in a British study (Neumann et al., 2009). Interestingly, N370S is rarely found in Asian populations (Mitsui et al., 2009), while L444P is found irrespective of ethnic background (Sidransky and Lopez, 2012).

Meanwhile, *GBA* is accepted as the most frequent genetic risk factor for Parkinson’s disease (Bras et al., 2015) with an OR of 5.43 (95% CI=3.89–7.57) (Sidransky et al., 2009) and our data suggest a possibly similar OR of 5.64 (95% CI=1.44-21.58) in dystonia. This makes it even more puzzling as to why the association between *GBA* and dystonia – like Parkinson’s disease – has evaded multiple large-scale attempts from epidemiologists and geneticists before. It has been argued that for Parkinson’s disease this might be due to a) epidemiologically, Gaucher’s disease being much rarer than Parkinson’s disease in the general population, b) genetic linkage studies not being able to detect this association as the *GBA* mutations are too rare in most but high-risk populations, such as Ashkenazy Jews, and c) genome wide association studies correcting for multiple-testing being too strict for this association to be detected, and also relying on the assumption that there is a single disease-associated allele per locus, which is not true for *GBA* with its multiple mutations (Rogaeva and Hardy, 2008). It appears possible that in addition to less large-scale genetic studies undertaken in dystonia than Parkinson’s disease, similar factors might have prevented the detection of *GBA* as a genetic risk factor in dystonia so far.

Third) the finding of a region-specific decrease in lysosomal function in human brain tissue in dystonia. So far, human post-mortem brain tissue has been assessed for lysosomal enzyme activity mainly in the context of Parkinson’s disease (Chiasserini et al., 2015; Gegg et al., 2012; Moors et al., 2019; Rocha et al., 2015). Apart from one study (Rocha et al., 2015), GCase activity has been consistently reported as decreased in the SN (Chiasserini et al., 2015; Gegg et al., 2012; Moors et al., 2019), while there is less agreement on decreased activity in cerebellum (Gegg et al., 2012; Rocha et al., 2015), putamen (Gegg et al., 2012), caudate (Chiasserini et al., 2015) or hippocampus (Rocha et al., 2015). Physiologically, GCase activity decreases with advancing age in SN and putamen (Rocha et al., 2015). Mutations in *GBA* are known to lead to reduced GCase activity, which has been semi-quantified in blood for L444P (<5% residual wild type GCase activity) (Grabowski, 2008), and N370K (17.7%) as well as E326K mutations (38.7%) in heterologous expression systems (Horowitz et al., 2011). There is so far only rudimentary understanding however, why enzyme activities vary even between cases with the same *GBA* mutation, although genetic modifiers, such as scavenger receptor class B member 2 (SCARB2) – encoding lysosomal integral membrane protein type 2 (LIMP-2), the receptor involved in the trafficking of GCase within the cell – or transcription factor EB (TFEB), regulating expression of a wider lysosomal gene expression network, respectively GCase activators, such as Saposin C, have been shown to modulate GCase activity (Siebert et al., 2014).

The brain regions studied here were chosen based on their presumed role in dystonia pathophysiology due to cerebellar (Kaji et al., 2018; Shakkottai et al., 2016), tectal (Holmes et al., 2012; Mc Govern et al., 2017) or basal ganglia dysfunction (Goto et al., 2005; 2013; Hanssen et al., 2018; Neychev et al., 2011), and tissue availability. In line with previous reports (Chiasserini et al., 2015), our healthy control data indicate enzyme- and brain region-specific activity levels for GCase as well as for b-GAL. Even with this relatively small sample, we observed an attenuation of this pattern in dystonia with an increase in the inter-individual variability of activity levels mainly in cerebellar structures. Using GCase activity as the main read-out and b-GAL as a lysosomal control enzyme, our data point at primarily cerebellar changes in this patient population. The detection of decreased GCase activity in cerebellar dentate nucleus adds significantly to the abnormal enzyme activity finding in peripheral blood, as it implies a systemic deficiency associated with dystonia. It is also in keeping with mounting evidence for a pathophysiological role of the cerebellum in dystonia (Filip et al., 2017; Neychev et al., 2011; Sadnicka et al., 2012; Shakkottai et al., 2016). Even further, it appears to point at a specific functional abnormality in the dentate nucleus, the main cerebellar efferent structure.

The above observations of decreased enzyme activity in central and peripheral tissue and genetics lend support to the hypothesis of lysosomal dysfunction in dystonia. Interestingly, in Parkinson’s disease decreased GCase activity has consistently been reported both in blood and brain (SN) of patients with (PD+GBA) (Alcalay et al., 2015; Atashrazm et al., 2018) and without *GBA* mutations (PD-GBA) (Gegg et al., 2012; Rocha et al., 2015). Similarly, mitochondrial Complex 1 deficiency (Mizuno et al., 1989; Schapira et al., 1989) has been found in the SN of Parkinson’s disease patients with (Morais et al., 2014) and without mitochondrial mutations (Flønes et al., 2018). Lysosomal dysfunction and Complex 1, i.e. mitochondrial deficiency is meanwhile acknowledged as a key pathophysiological mechanism in Parkinson’s disease (Poewe et al., 2017). Our finding of decreased lysosomal enzyme activity irrespective of *GBA* mutation status appears to follow the pattern of Complex I deficiency and lysosomal dysfunction in PD, pointing at a wider pathophysiological role of lysosomal dysfunction in dystonia. While the results presented in this work establish an association between lysosomal dysfunction and dystonia, they cannot prove a causal effect. The genetic analyses support the latter, though mutations in *GBA* seem to act as a modifier and risk factor and not in a mendelian fashion. Replication of our findings in other cohorts will be necessary to establish GBA mutations as a cause for dystonia. Given the relatively small sample size, genotype-phenotype correlation is currently not warranted.

The pathophysiology of dystonia as a syndrome is believed to primarily involve cerebral cortex, basal ganglia and cerebellum (Hanssen et al., 2018; Kaji et al., 2018; Neychev et al., 2011). So far, dystonia has been associated with lysosomal dysfunction as part of the clinical presentation in lysosomal storage disorders, such as chronic forms of GM1 gangliosidosis (Arash-Kaps et al., 2019), GM2 gangliosidosis (Meek et al., 1984), and less frequently in Niemann Pick type C and Kuf’s disease (Ebrahimi-Fakhari et al., 2018; Sedel et al., 2008). Human post-mortem neuropathology of the above disorders is scarce, but neuronal loss and gliosis primarily affecting striatum and pallidum has been reported in juvenile-onset, chronic GM1 gangliosidosis with dystonia (Suzuki, 1991), while selective neurodegeneration of cerebellar Purkinje cells has been reported in Niemann Pick type C (Walkley and Suzuki, 2004). Interestingly, GCase expression and enzyme activity is decreased in both PD+GBA and PD-GBA not only in SN but also cerebellum, a structure not primarily involved in Parkinson’s disease pathogenesis (Wu and Hallett, 2013). The fact that GCase activity in our samples was primarily affected in the cerebellum strengthens the role of the cerebellum in dystonia pathophysiology and points at a cerebellar lysosomal vulnerability, possibly shared by Parkinson’s disease and dystonia.

Given the above described recurrent overlaps and similarities between dystonia and PD regarding frequency and type of *GBA* mutations, peripheral and cerebellar enzyme activity pattern, it is important to carefully evaluate possible alternative interpretations of the above results. In patients presenting with dystonia in adulthood, particularly if symptoms start in the limbs, YOPD is an obvious differential diagnosis. Around 20% of YOPD patients may initially present with dystonia of variable degree, which might become even more prevalent with advancing disease (Wickremaratchi et al., 2011). The fact that, on careful clinical follow-up, we identified three YOPD cases (see Table 4) is in keeping with this notion. The exclusion of nigro-striatal degeneration in an additional, relevant proportion (16.4%) of dystonia cases supports the interpretation that these cases are unlikely to develop nigro-striatal degeneration. DaT scans had been ordered based on clinical context, i.e. suspicion of bradykinesia, or because of a suggestive family history, and although the evaluation of nigrostriatal degeneration hence was not done systematically in all participants, it was done systematically in suspicious cases. Furthermore, the time course between symptom onset and DaT scan argues against Parkinson’s disease underlying a dystonic presentation, as on average, time to diagnosis in YOPD – in keeping with the literature (Rana et al., 2012) – was 2.3 ± 0.5 years, while the mean disease duration to DaT scan in dystonia was 14.6 ± 11.8 years, making a (late) conversion highly unlikely.

For the large majority of cases in this series we are hence confident that we excluded an alternative underlying aetiology by exhausting differential diagnostic testing. However, the three cases that developed Parkinson’s disease with a latency of 14.0 ± 6.5 years after an initial diagnosis of cervical dystonia – two of which had a positive family history for cervical dystonia – warrant attention, as they raise the possibility that there might be a (rare) continuum between dystonia and Parkinson’s disease beyond YOPD. In this context, interestingly, olfactory dysfunction, a well-known and prevalent non-motor symptom in Parkinson’s disease, has recently been described as a potential novel endophenotype in cervical dystonia (Marek et al., 2018).

Limitations to our work predominantly apply to the heterogeneous, retrospectively collected cohort of combined dystonia cases: recruitment was performed at a tertiary referral centre’s movement disorder department, most certainly introducing selection bias. Retrospective data collection and the fact that genetic evaluations were not done by standardized means, i.e. a certain sequencing technique or gene panel, and that metabolic tests beyond lysosomal enzyme activities were done on a case-by-case basis, are inherent to such case series, but introduce additional heterogeneity and bias. Similarly, structural (CT, MRI) and functional (DaT scan) imaging was ordered on the basis of pre-existing documentation and individual presentation. On the other hand, the heterogeneous nature of cases included in this cohort in clinical practice precludes a “one-size-fits-it-all” diagnostic approach due to the multitude of theoretically possible aetiologies. The carefully chosen additional diagnostic tests hence reflect the individual clinical presentation. With this in mind, we can confidently exclude most common aetiologies, but possibly cannot exclude additional cases with a rare alternative aetiology of dystonia. Importantly however, this does not impinge on the argument for lysosomal dysfunction in dystonia.

Peculiarly, dystonia has only rarely been described as part of the clinical spectrum in Gaucher’s disease so far (Machaczka et al., 2018), which might be in part due to the scarcity of Gaucher’s disease itself and the clinical difficulty to detect subtle dystonic features, leading to underreporting (Ebrahimi-Fakhari et al., 2018). Alternatively, dystonia due to lysosomal dysfunction might be distinct from clinical Gaucher’s disease. The pattern of lysosomal activity differing from Gaucher’s disease, the fact that no obvious hepatosplenomegaly has been reported in any of the patients in this series, and the neuropathology pattern in Gaucher’s disease not affecting any of the brain structures classically involved in dystonia (Wong et al., 2004), argues for this.

The late conversion of cervical dystonia to Parkinson’s disease has not been reported so far and opens the possibility of a rare continuum between the two conditions. Further studies will have to establish a causal link, in particularly in showing in how far this occurs at a rate above the background, population and age-corrected incidence for Parkinson’s disease.

Isolated cervical dystonia constitutes a different clinical entity from combined dystonia and we want to avoid the impression that we would lump the two together. Nevertheless, the high proportion of *dystonia of unknown origin* among cervical dystonia and the identification of a similar metabolic pattern and *GBA* mutations in both groups argues for lysosomal dysfunction in dystonia as an overarching mechanism. As the identification of the disease mechanism has led to the successful establishment of enzyme replacement (Weinreb et al., 2002) and substrate avoidance therapy for Gaucher’s disease (Bennett and Mohan, 2013), the identification of lysosomal dysfunction in dystonia might similarly pave the way for causative therapeutic options in the future.

In summary, through comprehensive clinical, peripheral and central enzyme activity measurements and genetic analysis, this report provides first evidence for a role of lysosomal dysfunction in dystonia pathophysiology. Additional studies are needed to examine co-factors determining GCase activity beyond *GBA* genotype. Although the pattern of decreased lysosomal activity in the brain points at a possible role of the cerebellar dentate nucleus, the mechanism by which lysosomal dysfunction leads to a dystonic phenotype, remains to be established.

## Data Availability

Individual level data are available upon request from the authors.

## Abbreviations (alphabetical order)

b-GAL: beta-Galactosidase
CHIT: Chitotriosidase
CDN: Cerebellar dentate nucleus
CRB: cerebellar cortex
CoV: cerebellar cortex
GCase: Glucocerebrosidase
*GBA*: glucosidase acid beta
GBAP: glucosidase acid beta pseudo-gene
PALL: Pallidum
SCol: superior colliculus
wbc: white blood cell
YOPD: young-onset Parkinson’s disease

## Acknowledgments

The Queen Square Brain Bank is supported by the Reta Lila Weston Institute of Neurological Studies, UCL Queen Square Institute of Neurology.

## Supplementary figures and tables

**Figure S1:**
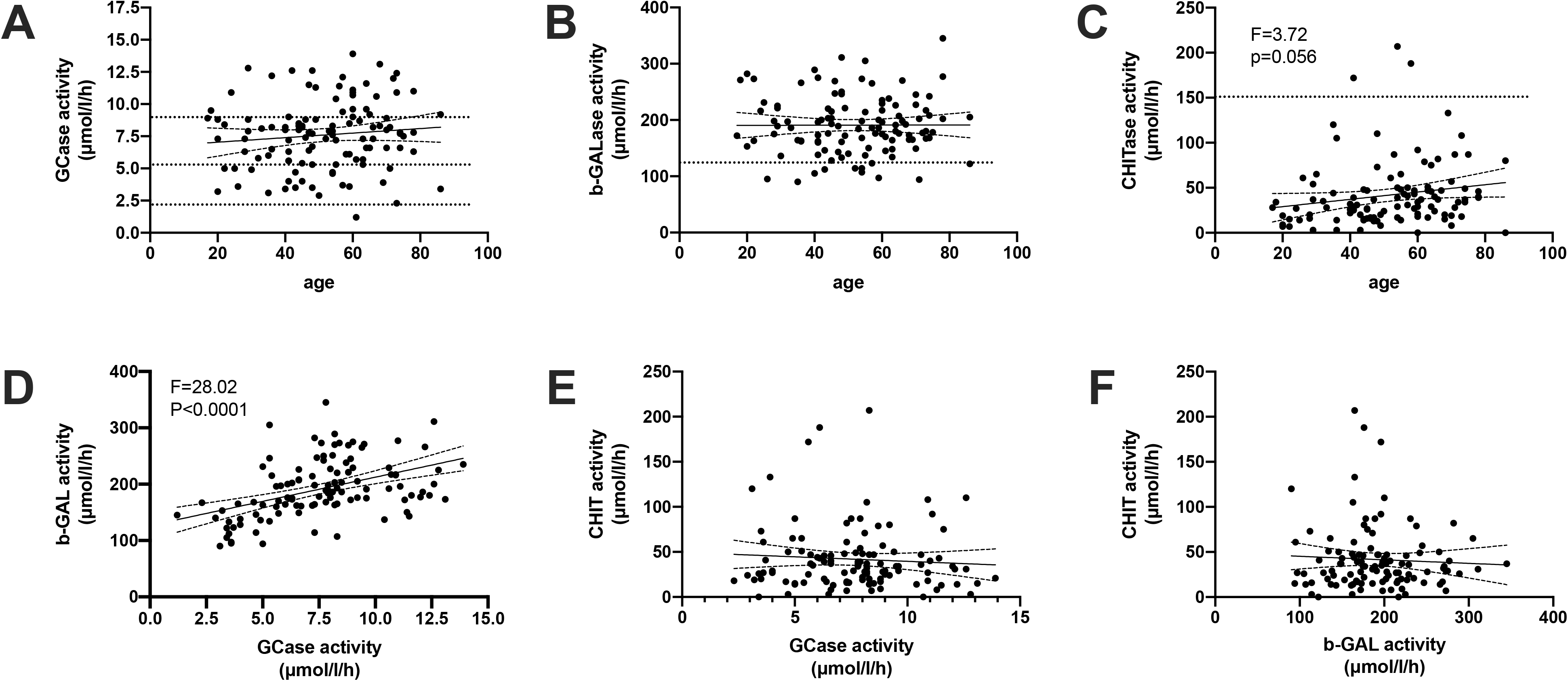
Correlation analysis of white blood cell lysosomal enzyme activity (in μmol/l/h) measurements. While there was no detectable correlation between GCase and b-GAL with age, CHIT activity showed a trend for higher activity levels with advanced age (P=0.056). Among enzyme activity measurements, there was a robust positive correlation between GCase and b-GAL (P<0.0001) but neither between GCase and CHIT (P=0.45) nor b-GAL and CHIT (P=0.57). Dotted lines indicate internal enzyme activity reference ranges – GCase: typical for homozygous *GBA* mutations (<2.4μmol/l/h), heterozygous *GBA* mutations (2.5-5.4μmol/l/h), heterozygous / unaffected overlap (5.4-8.9μmol/l/h) or unaffected (>8.9μmol/l/h); b-GAL: normal (130-303μmol/l/h); CHIT: normal (0-150μmol/l/h).

**Figure S2:**
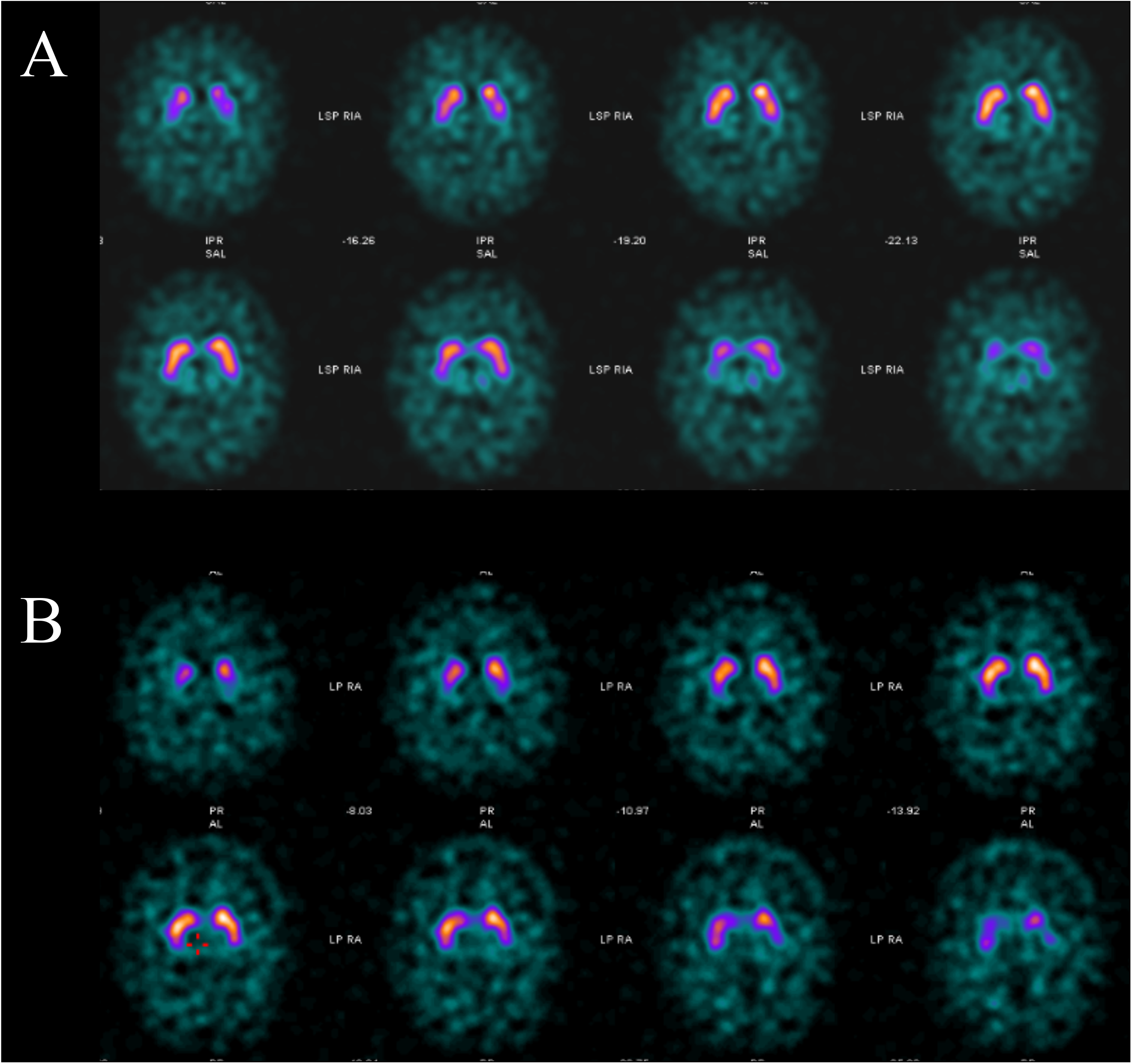
Serial negative DaT scans of case no.1 with identified *GBA* heterozygous N370S + T369M genotype taken 10 years (A) and 13 years (B) after dystonia onset.

### Supplementary material

#### Supplemental methods

##### Sanger Sequencing

In short, in a first step *GBA* was amplified in three fragments, spanning Exon1-5, Exon5-7 and Exon8-11 of the amplicon length. The PCR product length was measured in all samples using 1.5% Agarose gel electrophoresis to check if it was consistent with amplification of *GBA* (NM_000157.4, transcript ID: ENST00000368373.8; GBA-202;_GRCH38.p12 built) but not *GBAP*. In a second step, individual exons were sequenced. For the amplification PCR, 7.5μl fast start PCR master mix (Roche no.10368820), 0.5μl of 10μM forward primer, 0.5μl of 10μM reverse primer, 5μl deionised water and 1.5μl genomic DNA template (at a concentration of 40-80ng/μl) were used, resulting in a total volume of 15μl per reaction. After ExosAP cleaning of the PCR product (50μl Exonuclease I (Thermo Fisher no. EF0651), 200μl FastAP (Thermo Fisher no.EN0582) and 750μl deionised water mixed with the PCR product at a ratio of 2:5), 3μl ExosAP product was mixed with 3.5μl deionised water, 2.0μl Sequencing Buffer and 0.5μl BigDye Terminator v1.1 (both Applied Biosystems, no 4336699) and 1.0μl primer and sequencing performed on a 3730 DNA Analyzer (Thermo Fisher, no. 3730S).

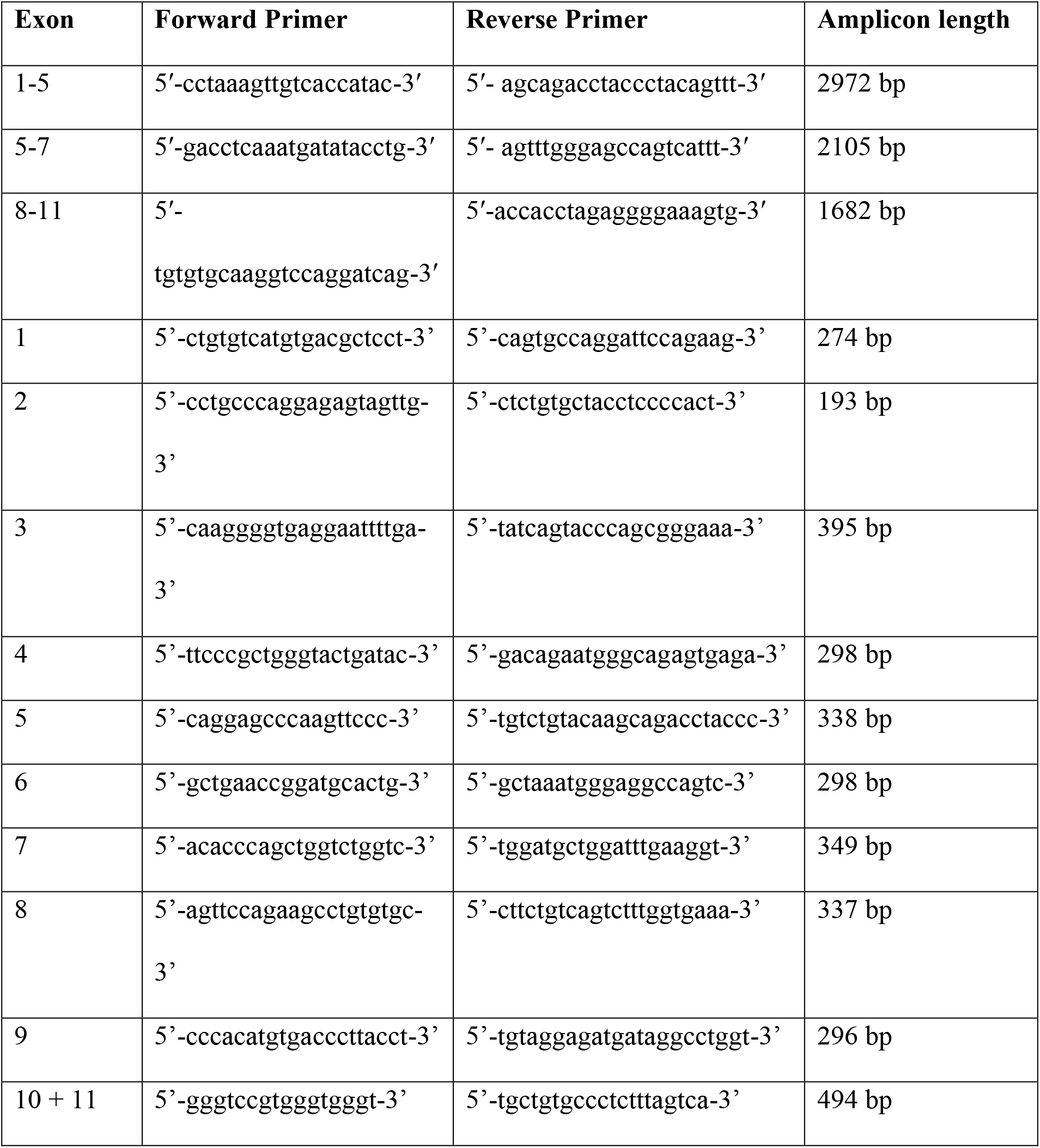

Summary of primers used for *GBA* sequencing including their 5’-to-3’ nucleotide sequence and resulting amplicon length.

The PCR light cycler was programmed as follows for the amplification of *GBA* fragments:

1. PCR program for the amplification of *GBA* Exons 1-5

2. PCR program for the amplification of *GBA* Exons 5-7

| **Amplification PCR *GBA* Exon1-5** |  |  |  |  |  |  |
| --- | --- | --- | --- | --- | --- | --- |
| Time (min) | 15:00 | 00:45 | 00:45 | 03:00 | 07:00 | Hold |
| Temperature (°C) | 94.0 | 94.0 | 58.0 | 72.0 | 72.0 | 4.0 |
| n cycles | 1 | 37 |  |  | 1 |  |

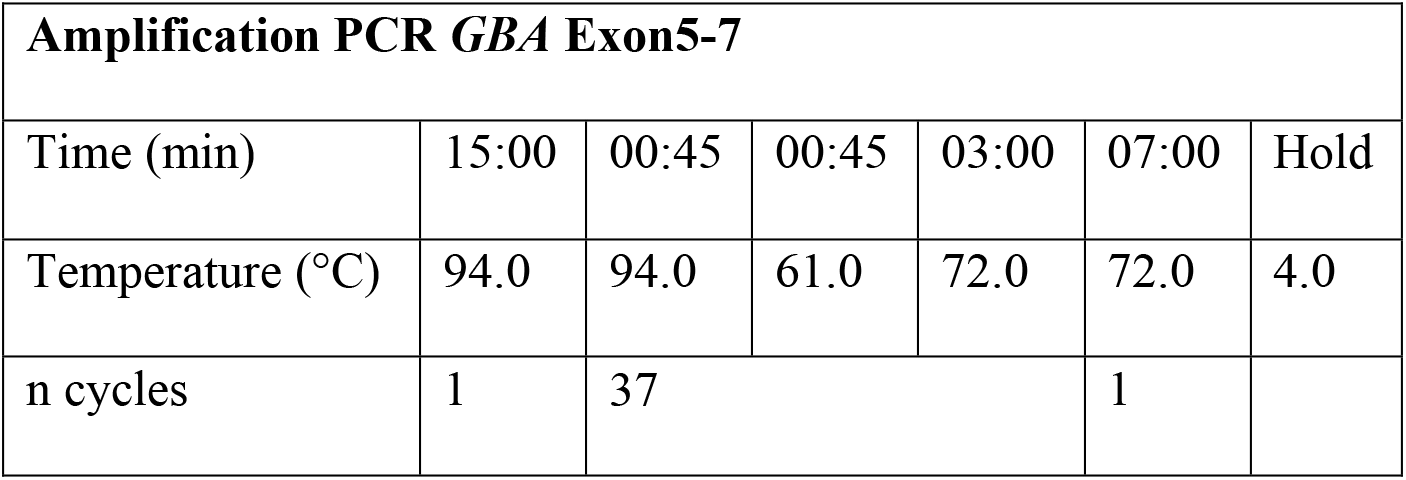
3. PCR program for the amplification of *GBA* Exons 8-11

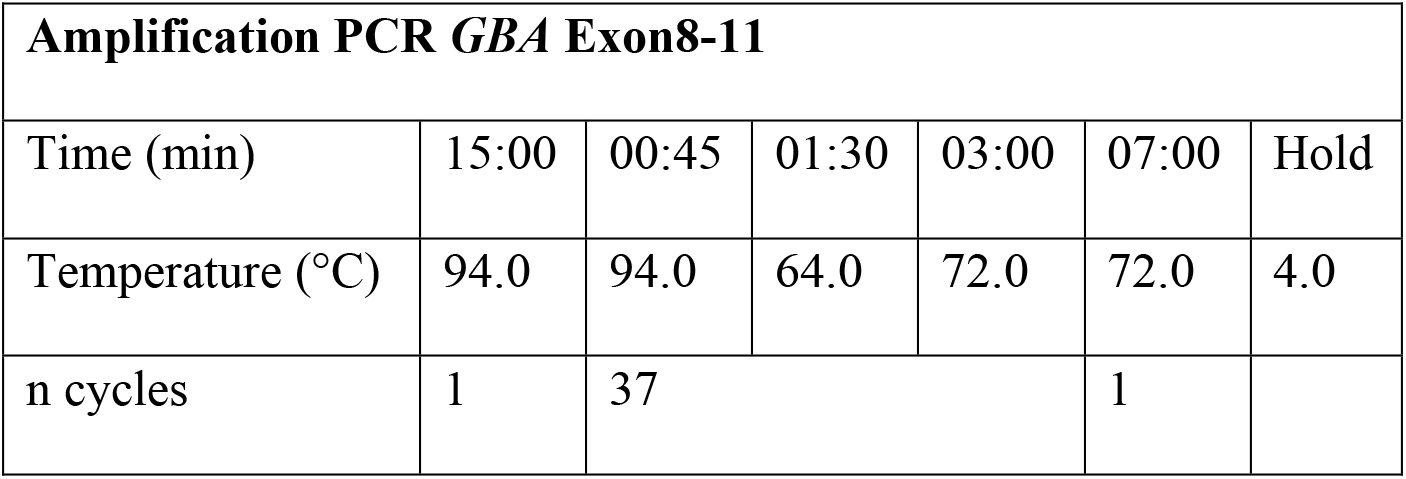

PCR program for the ExosAP cycle

| **ExosAP Cycle** |  |  |  |
| --- | --- | --- | --- |
| Time (min) | 30:00 | 15:00 | Hold |
| Temperature (°C) | 37.0 | 80.0 | 4.0 |

PCR program for Sanger sequencing reaction

| **Sequencing 3730 cycle** |  |  |  |  |  |
| --- | --- | --- | --- | --- | --- |
| Time (min) | 01:00 | 00:30 | 00:15 | 04:00 | Hold |
| Temperature (°C) | 94.0 | 94.0 | 50.0 | 60.0 | 4.0 |
| n cycles | 1 | 25 |  |  |  |

#### Enzyme activity measurements

Protein concentration and enzyme activity measurements were performed according to internal laboratory protocols at the UKAS-accredited Clinical Enzymology Laboratory at Great Ormond Street Hospital.

For protein concentration measurements, Bovine Serum Albumin (BSA) (1 mg/ml in 0.15M NaCl and 0.05% sodium azide; Sigma P 0914) was used for standard curve calculation. Each sample was prepared in two concentrations (2.5μl sample + 47.5pl distilled H_2_0; 5μl sample + 45 μl distilled H_2_0) and the higher of both measurements used as the final protein concentration. After the addition of 1ml bicinchoninic acid (BCA) (Sigma B9643), each sample was incubated at 37°C for 10 minutes. After addition of 20μl of 4% Copper sulphate solution (Cupric sulphate, (CuSO_4_.5H_2_O) VWR 100913P), and 20min incubation at 37°C, absorbance was read at 562nm (Cecil 2040 Spectrophotometer).

The mean protein concentration in enzyme preparation was calculated from the standard curve (mg/ml). Duplicate protein concentrations outside a margin of error of 20% repeated.

For protein assay, samples stored fast frozen were diluted to a concentration of 2mg/ml. Eppendorf cups containing 80μl Na taurocholate solution (Na taurocholate pure (Calbiochem; no. 580218); 20mg/ml H2O, 37.2mmol/L) and 20μl enzyme solution containing 10-60μg protein per sample (20 μl water for blank samples) were prepared and 100μl substrate solution (4–methylumbelliferyl-β-D-glucopyranoside (MWt. 338) Melford M1097; 1.69 mg substrate / ml McIlvaine citrate-phosphate pH 5.4 buffer (5 mmol/L), dissolved by warming to 80°C) added at timed intervals and incubated at 37°C for 60minutes. At timed intervals 1.0ml stopping reagent (0.25 M glycine buffer pH 10.4) was added. Fluorescence was read on a Fluorimeter (Perkin Elmer LS 55) with the excitation set at 365nm at an emission wavelength of 450nm. The enzyme activity was calculated in nmol/hour/mg protein using the below formula: Average the test (T) and blank readings (B).

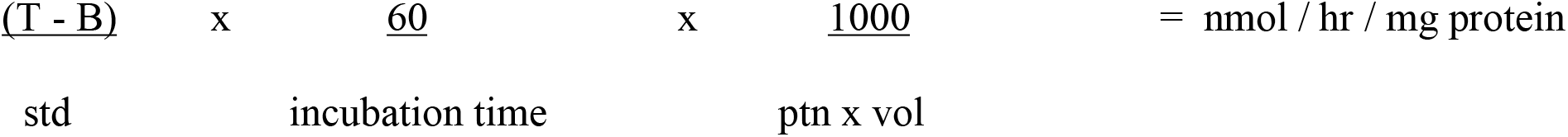

